# Survival of people with untreated tuberculosis: effects of time, geography and setting

**DOI:** 10.1101/2022.12.15.22283231

**Authors:** Carly A Rodriguez, Sarah V Leavitt, Tara C. Bouton, C Robert Horsburgh, Pia Abel zur Wiesch, Brooke Nichols, Helen E Jenkins, Laura F White

## Abstract

**Background:** An estimated 40% of people who developed tuberculosis in 2021 were not diagnosed or treated. Pre-chemotherapy era data are a rich resource on survival for people with untreated TB. We aimed to identify heterogeneities in these data to inform more precise use of them.

**Methods:** We extracted survival data from pre-chemotherapy era papers reporting TB specific mortality and/or natural recovery data. We used Bayesian parametric survival analysis to model the survival distribution, stratifying by geography (North America versus Europe), time (pre-1930 versus post-1930), and setting (sanitoria versus non-sanitoria).

**Results:** We found 12 studies with TB-specific mortality data. Ten-year survival was 69% in North America (95 CI: 54%-81%) and 36% in Europe (95% CI: 10%-71%). Only 38% (95% CI: 18%-63%) of non-sanitorium individuals survived to 10 years compared to 69% (95% CI: 41%-87%) of sanitoria/hospitalized patients. There were no significant differences between people diagnosed pre-1930 and post-1930 (five-year survival pre-1930: 65%; 95% CI: 44%-88% versus post-1930: 72%; 95% CI: 41%-94%).

**Conclusions:** Mortality and natural recovery risks vary substantially by location and setting. These heterogeneities need to be considered when using pre-chemotherapy data to make inferences about expected survival of people with undiagnosed TB.

## INTRODUCTION

Tuberculosis (TB) is a major infectious cause of global mortality, with case detection (an estimated 40% were undiagnosed in 2021) and failure to cure those treated being major drivers of mortality and transmission ^1^. Clinical outcomes of individuals with untreated TB are important from a public health perspective and in modelling studies.

Several studies have mined the rich pre-chemotherapy era TB literature to understand outcomes for people with untreated TB^2–4^. As we continue to use these important pre-chemotherapy era data, it is important to remember that these studies are not homogeneous. They span decades during which tumultuous events occurred (e.g. World Wars, the Great Depression), sanatoria were for treatment to varying degrees, studies were carried out in differing geographic regions (North America and Europe), and the quality and availability of medical care increased over time. TB outcomes among untreated individuals could vary significantly by these factors.

Here, we use studies from the pre-chemotherapy era that recorded mortality and natural recovery outcomes over time. We aim to estimate TB mortality rates during this era overall and by geographic and temporal context hypothesizing that individuals from countries with more resources during times of great stability would have better outcomes than individuals in studies from more turbulent times. We also aim to explore outcome differences between sanitoria patients versus those not in sanitoria and also hypothesize that individuals in sanitoria had less severe disease initially than those who were not.

## METHODS

### Search strategy

Since the development of anti-TB chemotherapy in the mid-twentieth century, studies reporting outcomes of untreated populations are unethical and pre-chemotherapy era studies are often not indexed in databases (e.g. PubMed, EMBASE). Therefore, we reviewed our personal libraries, which include publications of pre-chemotherapy era cohorts from a previous study of untreated children with TB ^4^, and publications used by ^2^ in a similar review to this current study (Table S1). We additionally reviewed the reference lists of these publications for further sources, and reference lists of those papers, where possible, to capture additional data on adults to supplement the original review results.

### Review of studies

We included cohort studies comprised of TB patients who did not receive anti-TB chemotherapy, and that reported the proportion of the cohort within specified time interval(s) with the following four outcomes: died from TB, died from a non-TB cause, naturally recovered, and lost to follow-up (Figure S1). Outcome definitions would depend on specific publications and are included in the results section and Supplement (Table S2). “Natural recovery” in this literature was not clinically well-defined and our definition would need to be driven by publications that we identify. However, natural recovery would likely include patients who no longer had detectable *Mycobacterium tuberculosis* (*Mtb)* in sputum smears, were able to work, or had symptom resolution ^5^.

We excluded publications that were not about TB, did not contain data on natural recovery or TB-specific mortality, were restricted to patients with specific forms of TB (e.g. TB meningitis) or narrow age groups (e.g. children <2 years old), case reports, autopsy studies, or were primarily patients receiving anti-TB drugs. We excluded publications presenting population-level outcome rates, or that did not specify follow-up time or loss to follow-up. Additionally, we excluded reviews or editorials but we did search their reference lists for additional studies of interest.

Two reviewers (two of TCB, HEJ, CAR, LFW) independently read each publication to determine inclusion eligibility. Non-English publications were translated by authors fluent in the relevant language (PAzW, BN) and independently reviewed by HEJ and LFW, along with the relevant translator, to determine eligibility. Discrepancies were resolved by group discussion.

### Data extraction

Two reviewers (two of TCB, HEJ, CAR, LFW, PAzW, BN) independently extracted data from each publication using a standardized electronic form. We extracted: study type (e.g. population-based cohort, sanatorium-based cohort), cohort characteristics (geographic location, time period, forms of TB disease, age range), determination of death, outcome definitions, treatment (e.g. sanatorium, surgery, anti-TB drugs), and the start of follow-up time (i.e. notification/diagnosis, sanatorium entry or exit). For consistency among sanatorium studies, if the follow-up start time was the sanatorium exit and the average sanatorium stay was reported, the follow-up times were shifted ahead by this average length of stay so that the follow-up start was approximately entry into the sanatorium. If the average length of stay was not reported, then the follow-up times were shifted by 165 days, the average length of sanatoria stay in the United States between 1934-1938 ^6^. We extracted data into life tables and stratified by disease severity when possible.

Publications often reported on multiple cohorts and occasionally, the same study reported data over multiple publications. Therefore, we enumerated findings using the following terminology: “publications” are individual published reports in the literature; “studies” are a population of patients recruited, diagnosed, and treated in a similar fashion during the same time period, which may be reported over multiple publications; and “cohorts” are a patient population that may have been recruited, diagnosed, and/or treated differently over varying time periods but were in the same publication.

Discrepancies between reviewers were resolved by group discussion and reconciled extractions were entered into REDCap electronic data capture tools hosted at Boston University Medical Campus, Boston, MA, USA. Ethics approval was not required because all data were from published reports.

### Mortality meta-analysis

We transformed the extracted life-table structured data and created individual-level data with the interval from entry/diagnosis to death or time of censoring for each individual in each cohort. We used parametric survival analysis with interval censoring to estimate the survival times for TB-specific mortality. TB deaths were considered events and deaths from other causes were considered censored observations.

We used a Bayesian framework to estimate the survival distribution which included a frailty term for each study (see Supplement). We analyzed all of the studies together in one model and then we compared mortality outcomes by time (pre-1930 to post-1930), geography (North America versus Europe) and setting (sanatorium/hospital studies and non-sanatorium studies) due to the difference in care pattern and start of follow-up time. We choose 1930 as a cut-off as the approximate mid-point of the total time period covered by our included studies. For all analyses, we plotted the study-level survival curves overlaid with the overall survival curves (the mean of the frailty distribution) with 95% credible intervals. We estimated study-specific one, five, and ten-year survival probabilities and median survival times with 95% credible intervals.

We compare the baseline reported severity in individuals treated in sanitoria/hospitals to those not treated in sanitoria, where disease severity reporting is available. We provide summary statistics of this data and test for associations using a chi square test.

We used R v4.0.2 with Bayesian models fit using JAGS with R2jags v0.6-1. Model diagnostic results are in the supplement. All data and code are available on GitHub at https://github.com/sarahleavitt/TB_mortality.

## RESULTS

### Literature review

We identified 153 publications (93 from personal libraries, 60 from a manual reference search of personal library publications) during screening and obtained 142 (93%) for full-text review (Figure 1). We excluded 122 publications, most due to not reporting an outcome of interest (n=48), unclear follow-up time (n=23), or reporting population-level rates (n=17). Data from 20 publications were included; three publications included data from the same cohort but were stratified by disease severity across three publications. For two studies ^7,8^, data required modification to conform to a life table format (see Supplement).

**Figure 1.**
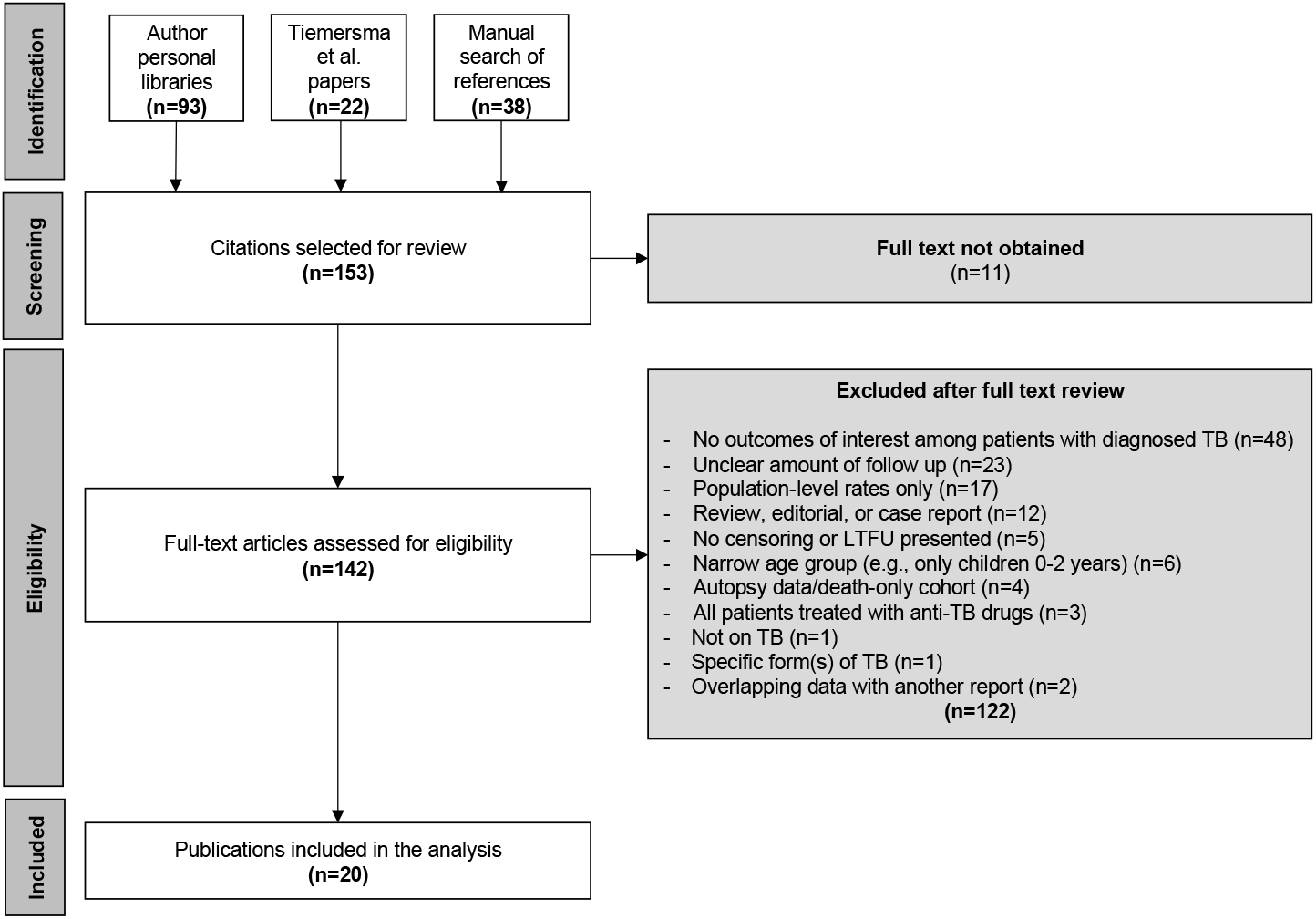
Flow chart describing the review process and exclusions.

### Characteristics of cohorts

Of the 20 included publications, 10 (50%) were from North America, 5 (25%) from the United Kingdom, 3 (15%) from Germany (one from an area that is now part of Poland), 1 (5%) from the Netherlands, and 1 (5%) from Norway (Table 1). Twelve studies (60%) were conducted primarily before 1930. Most publications enrolled patients between the 1910s and 1940s. Ten (50%) publications included patients treated at a sanatorium or an inpatient hospital. Six (30%) publications indicated a subset of patients received surgical treatment; anti-TB chemotherapy was provided to a minority in two publications (Table 1).

**Table 1.**
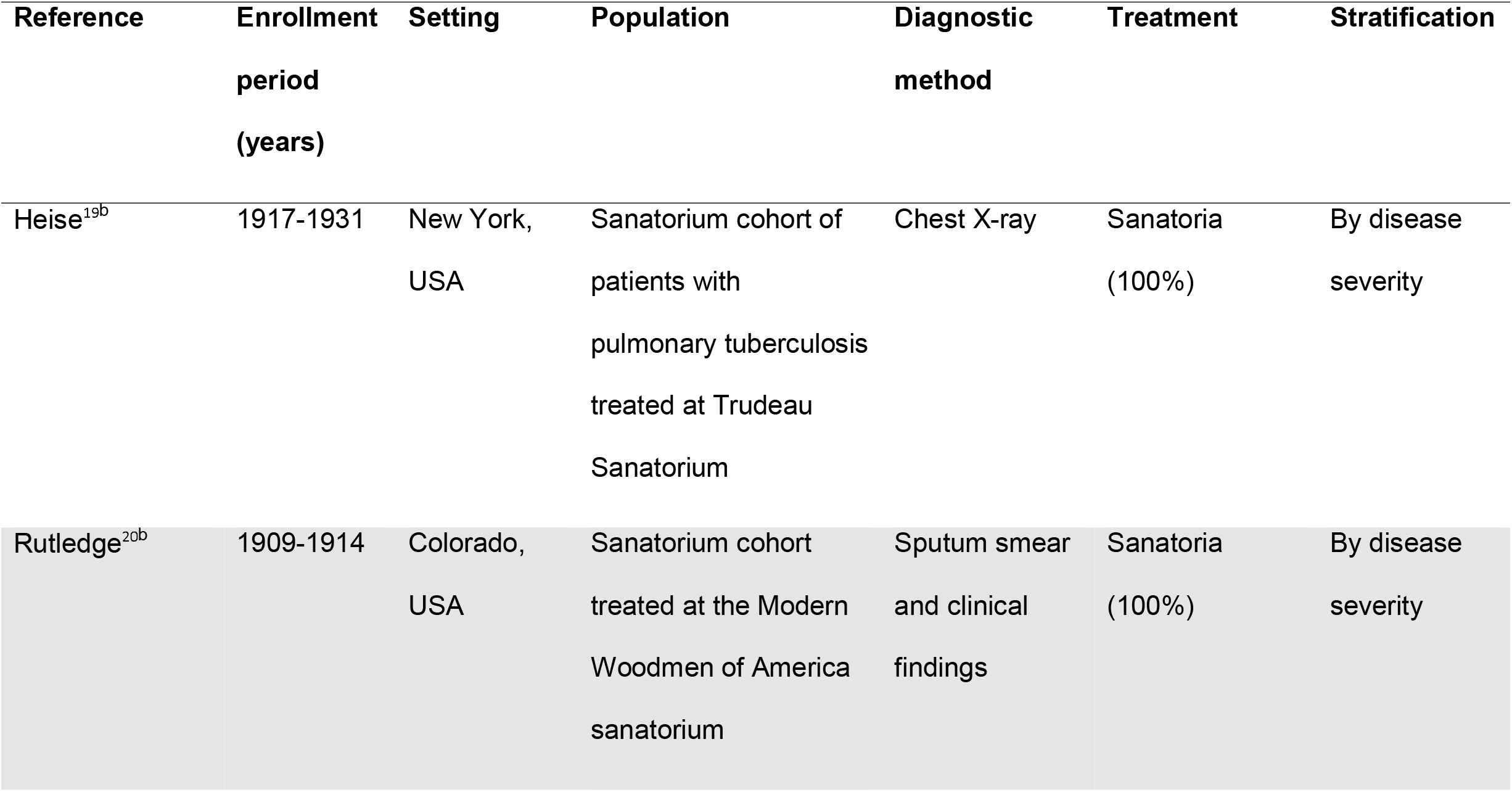
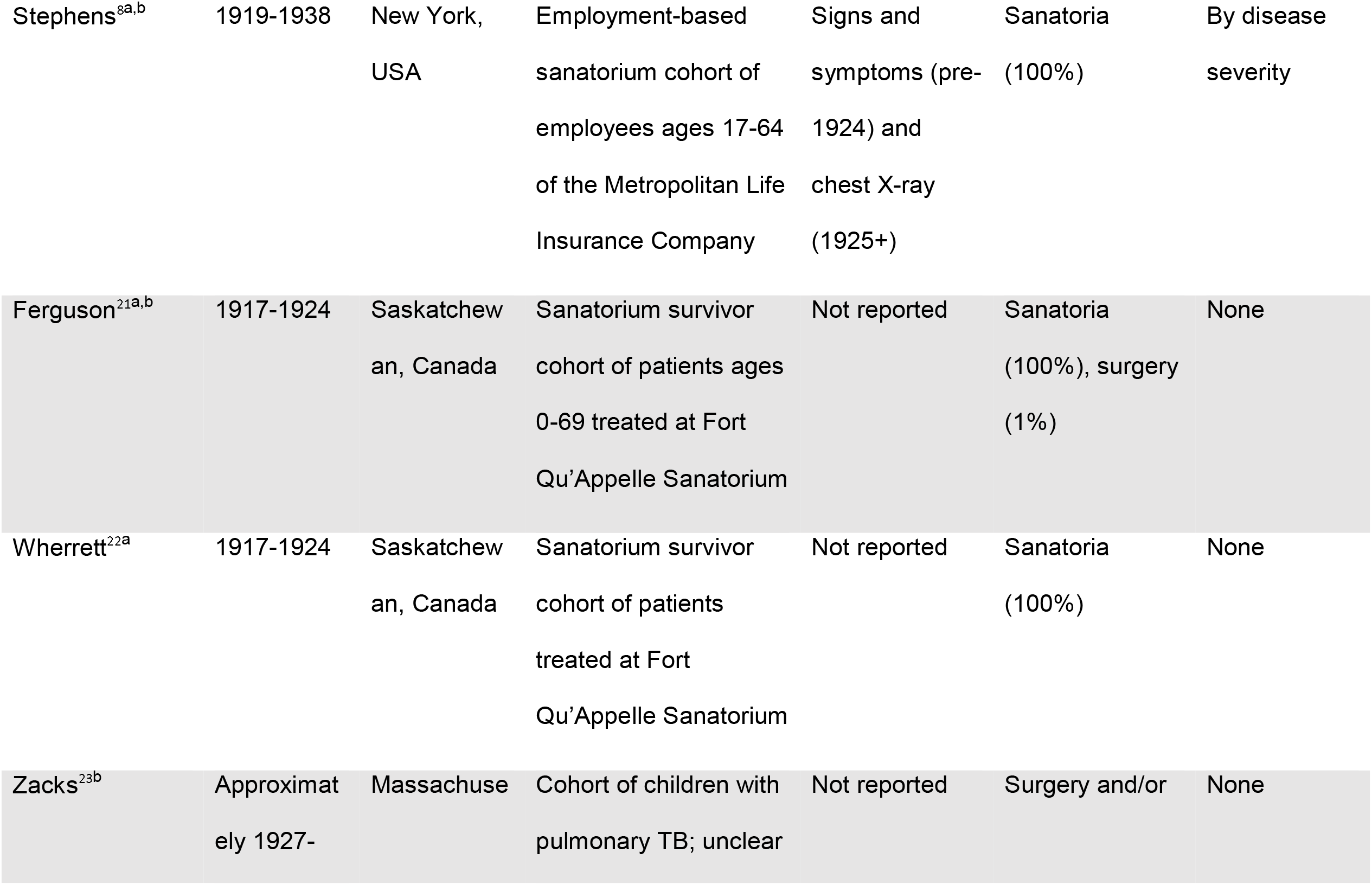
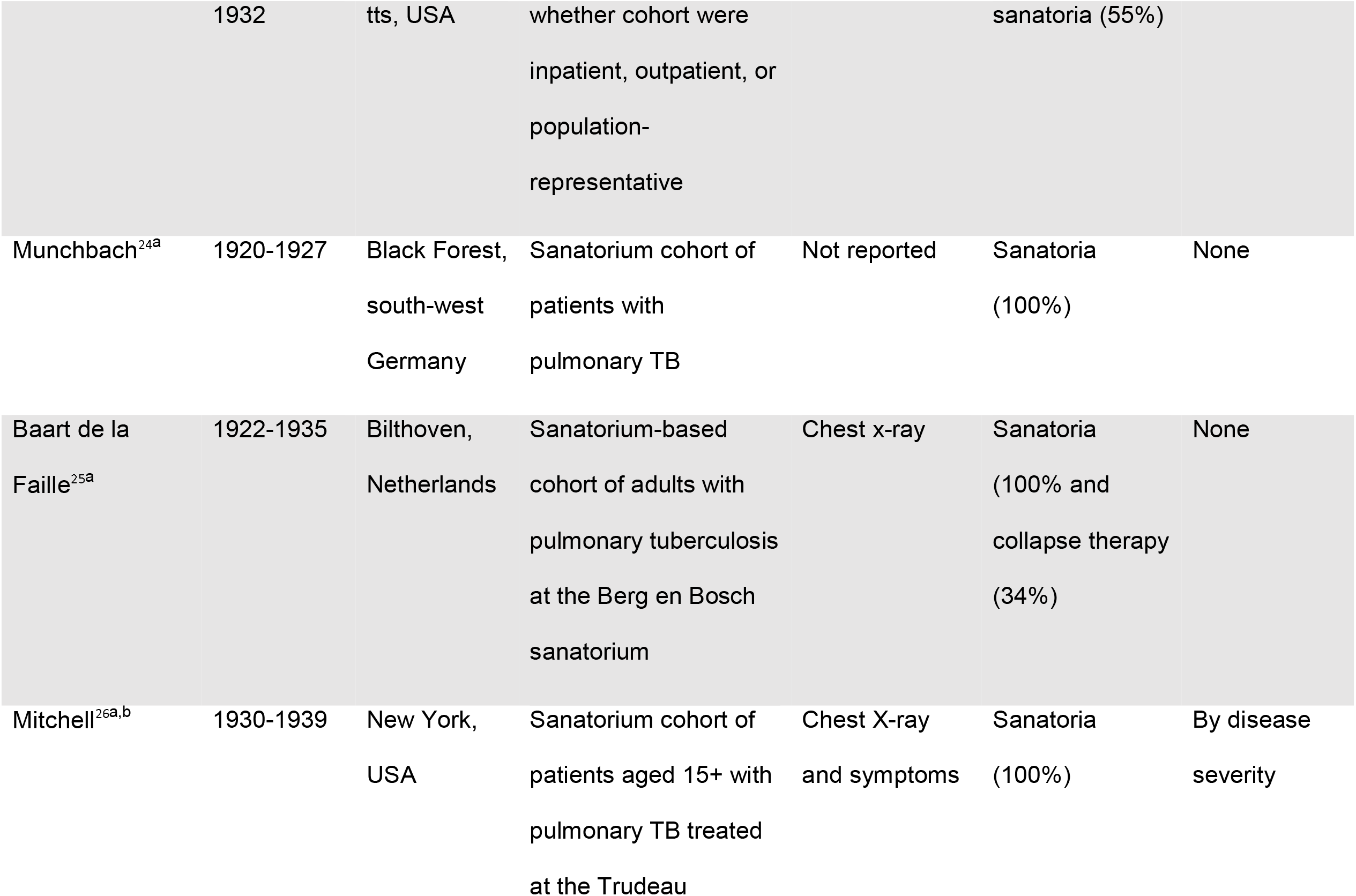
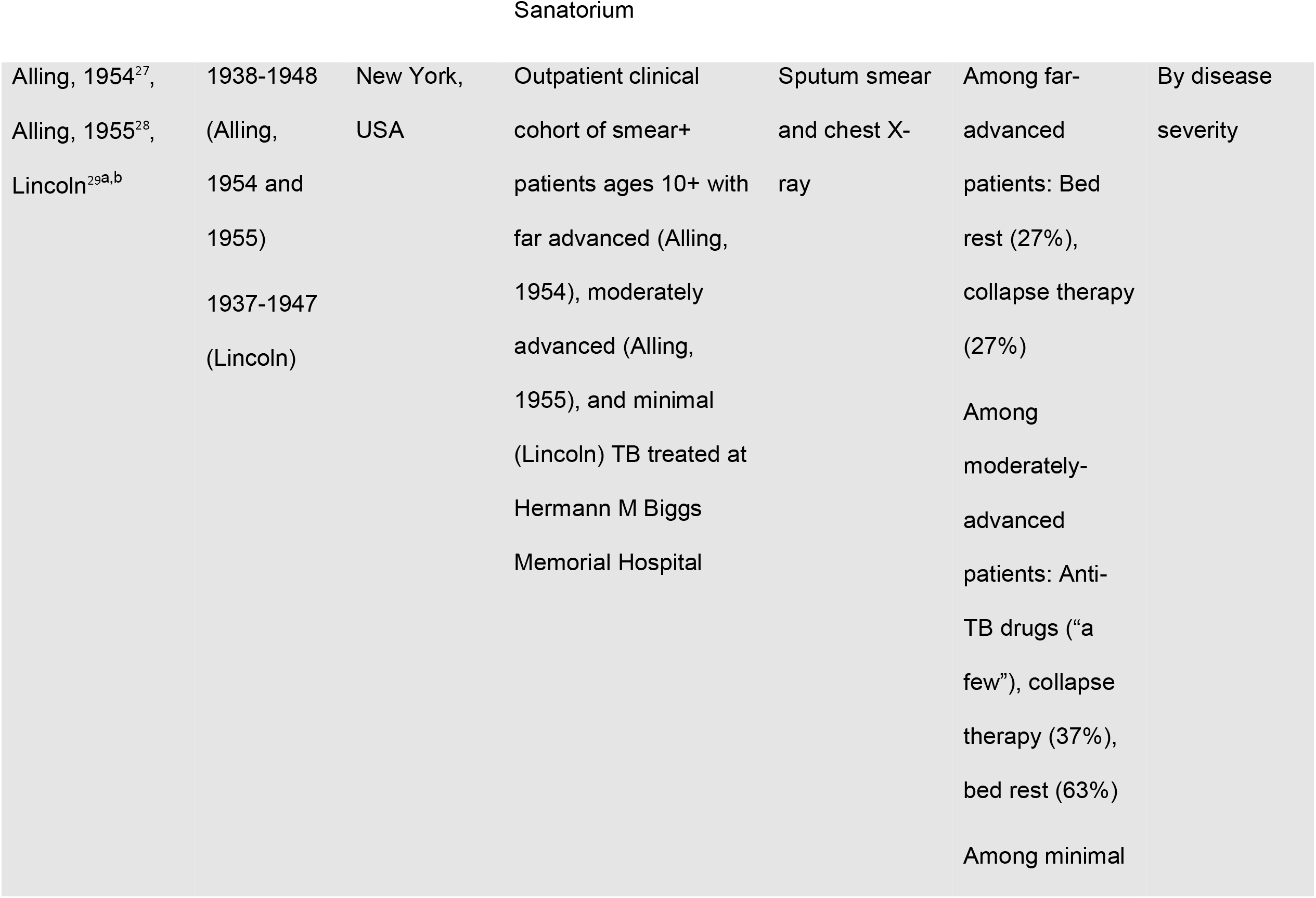
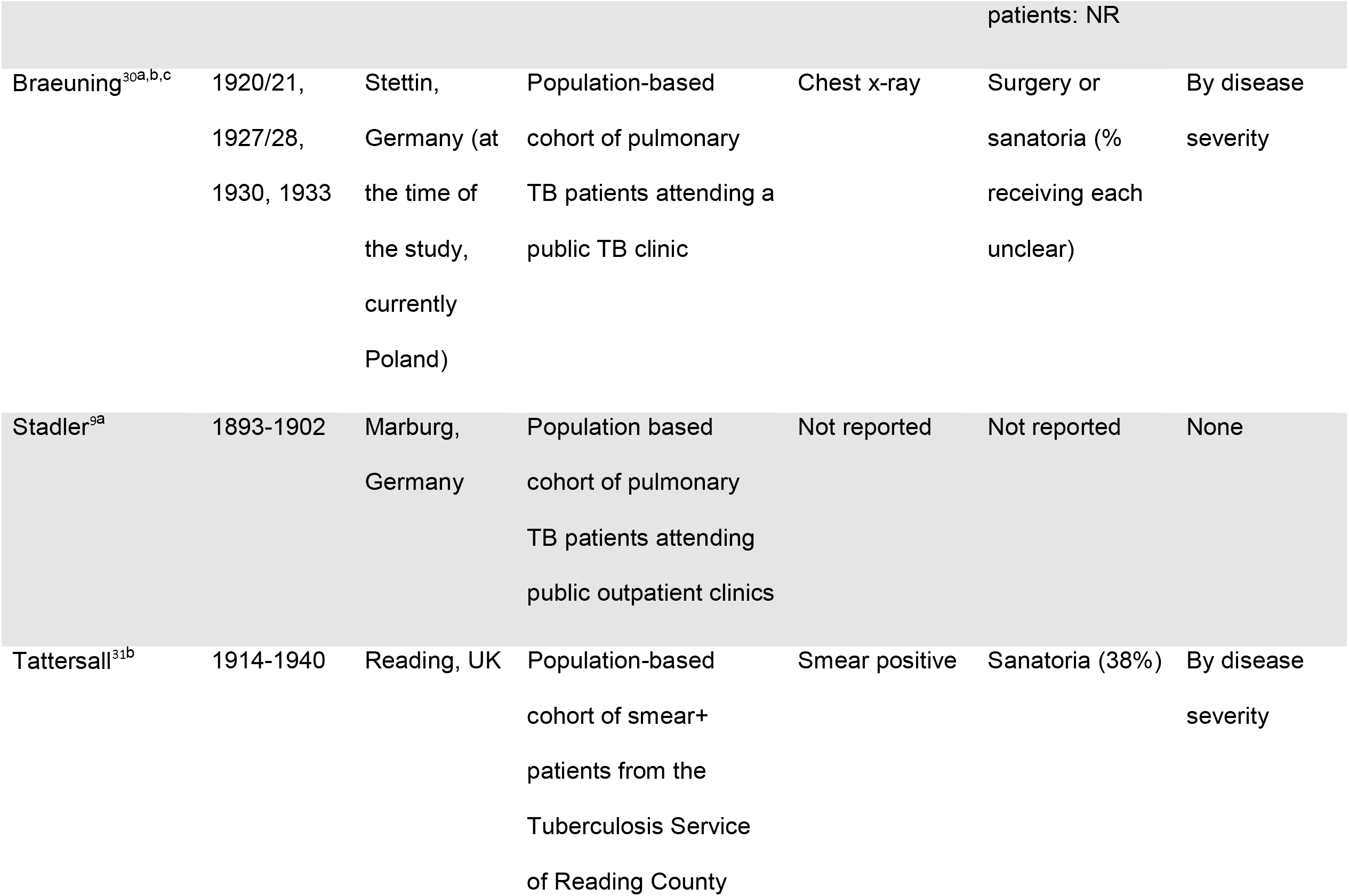
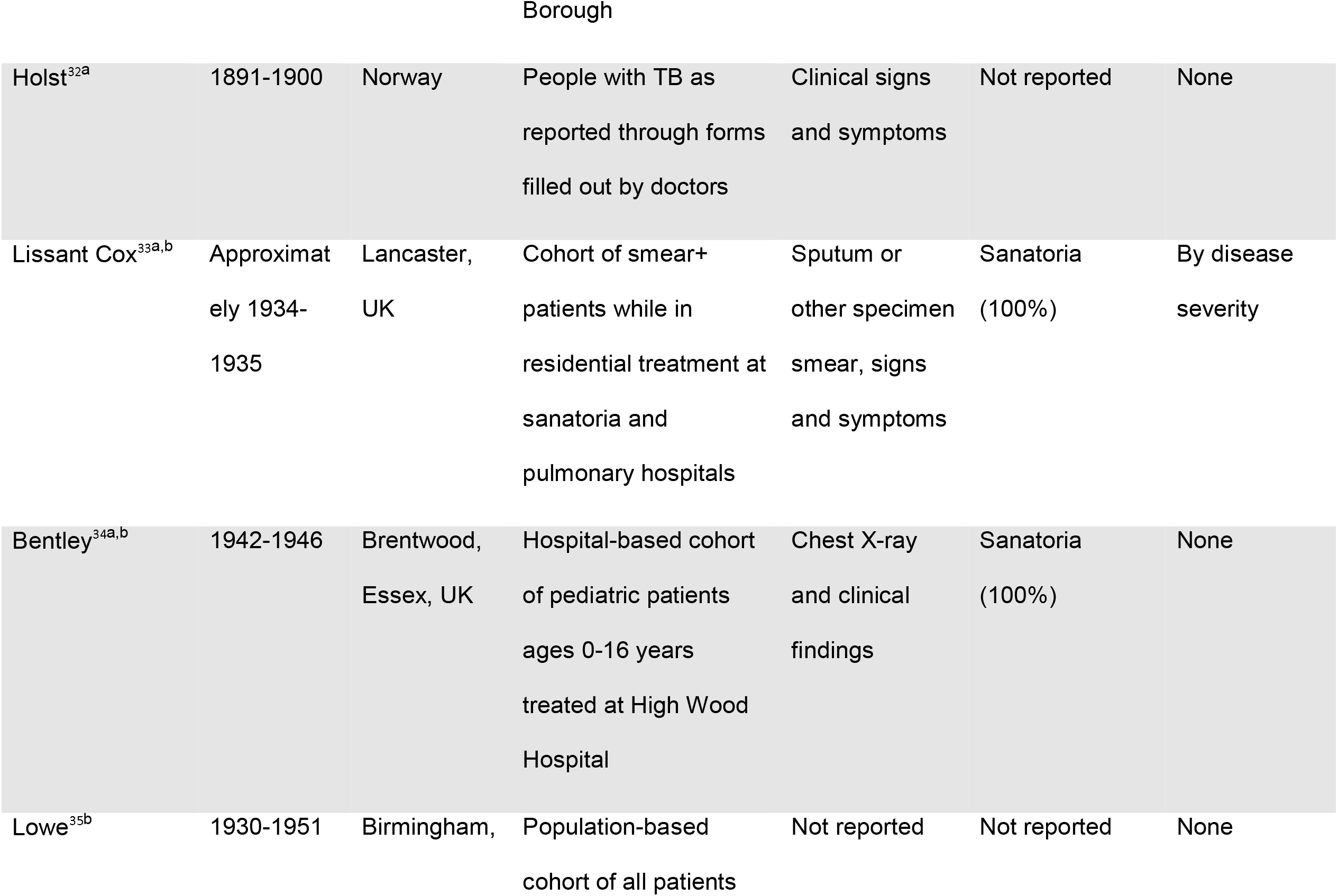
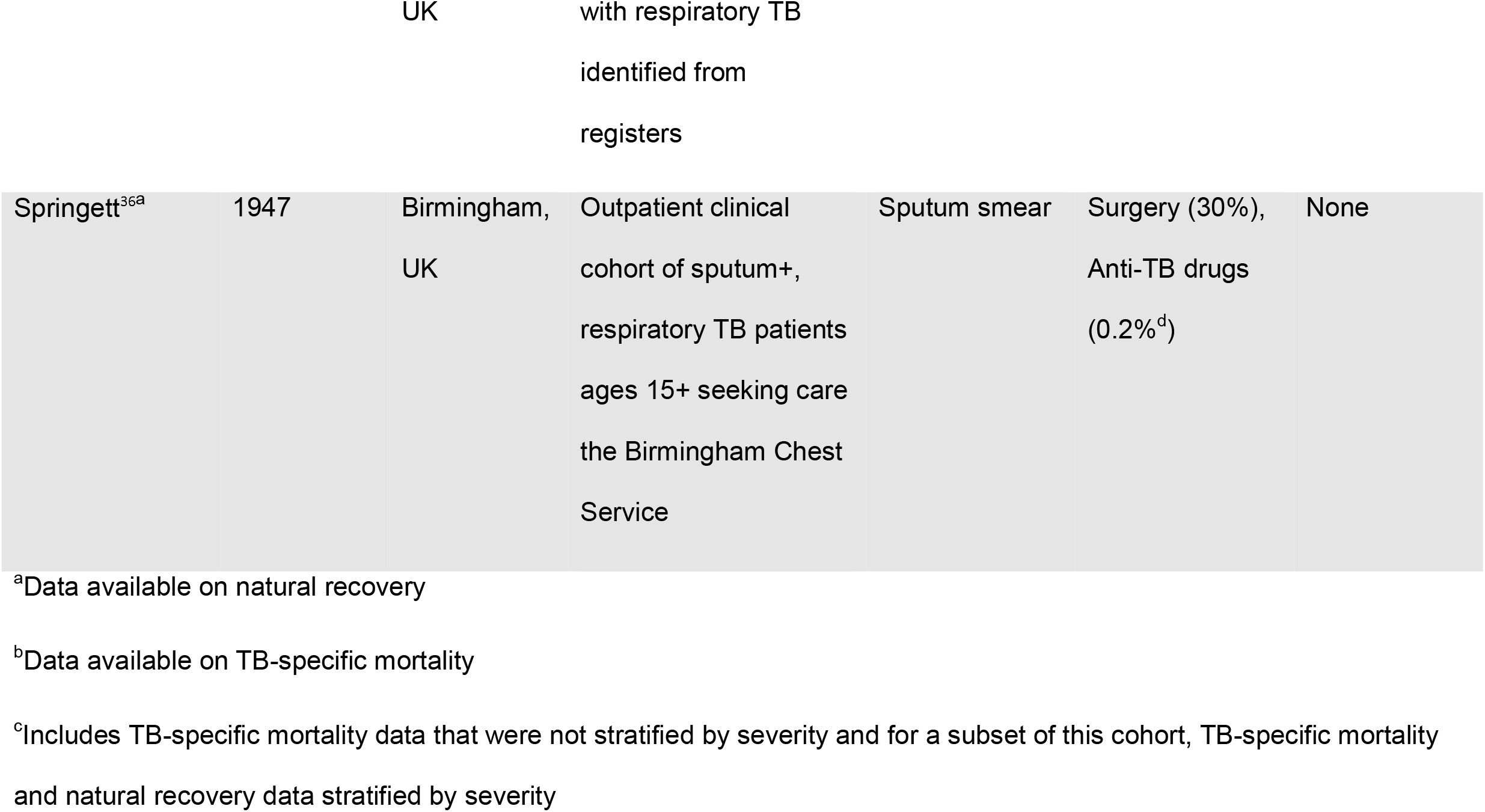
Descriptive characteristics of pre-treatment era cohorts with data on tuberculosis-specific mortality and/or, natural recovery from tuberculosis (N=20 publications, 18 studies)

Outcome definitions were not standardized during the pre-chemotherapy era and definition reporting was minimal in some publications (Table S2). Only five (25%) studies described determination of death. These included clinician-determined cause of death, via questionnaire mailed to households, or through multiple sources such as family report or vital record review. Natural recovery definitions included patients with no longer detectable *Mtb*, able to work, and the more stringent, standardized definitions of the United States’ National Tuberculosis Association’s (USNTA) *Diagnostic Standards*, which required symptom resolution, negative culture, and improved chest x-ray for ≥two years to establish recovery (Table S2) ^5^.

The 20 publications represented 18 studies. The 18 studies followed 84 cohorts with some studies following one cohort and others identifying different cohorts at each time point for a total of 35,900 patients. Of the 18 studies, 12 (67%) representing 53 (63%) of the cohorts reported TB-specific death. Of the 18 studies, 12 (67%) representing 63 (72%) of the cohorts classified alive patients as either having chronic TB or having naturally recovered by each time point.

### Overall mortality results

Using all data, there were 12 studies, 53 cohorts and 17,166 patients with follow-up data on TB-specific mortality. Assuming a lognormal distribution, we estimated TB-specific mortality survival (meanlog=2.51, sdlog=1.72). Overall, 93% of individuals survived one year (95% CI: 0.84-0.98) while 55% of individuals survived 10 years (95% CI: 0.36-0.73) with a median survival time of 12.33 years (95% CI: 5.47-28.93) (Table 2, Figure S3). Model fit diagnostic plots for all models are in the supplement (Figures S11-S17).

**Table 2.**
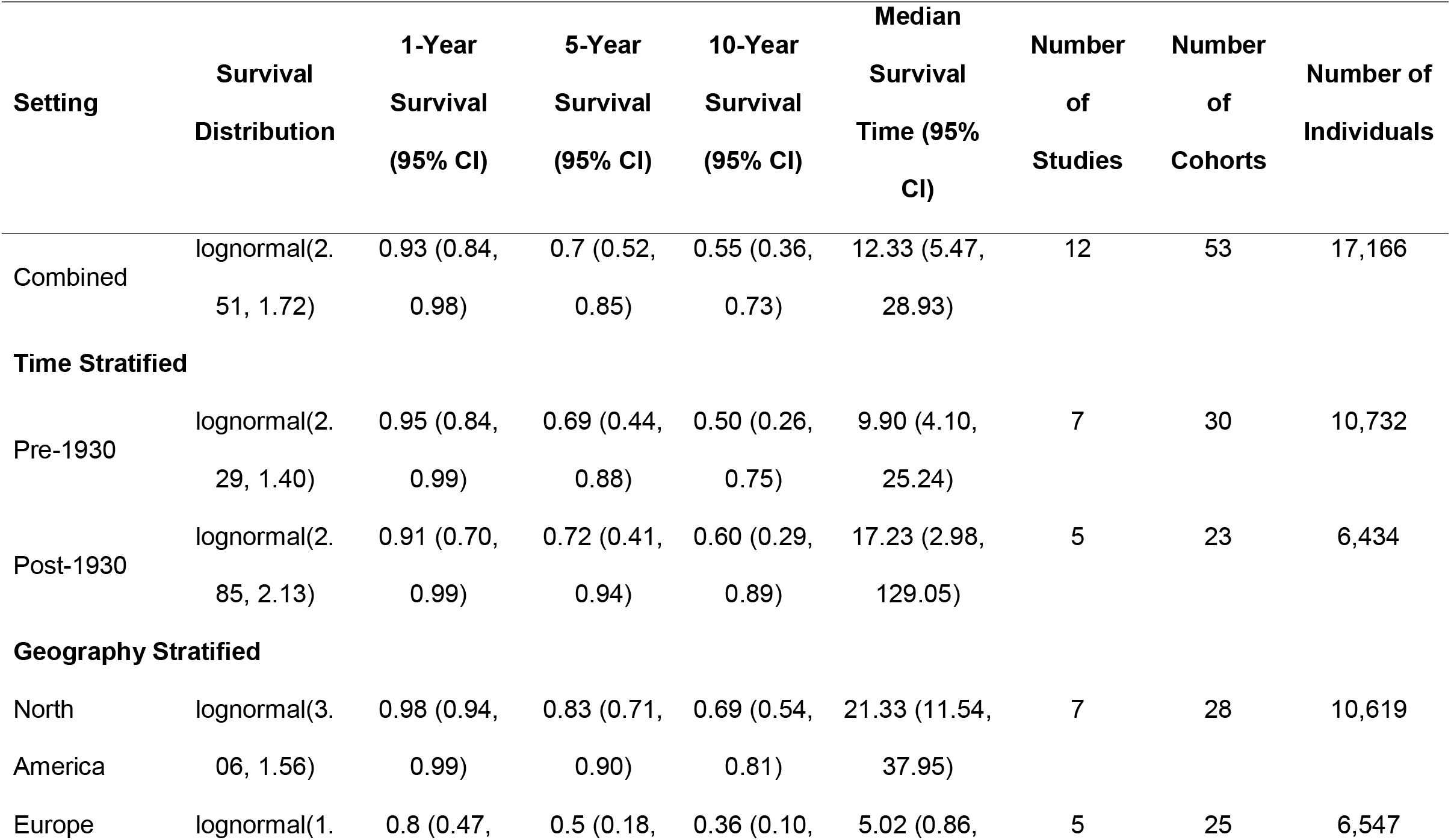
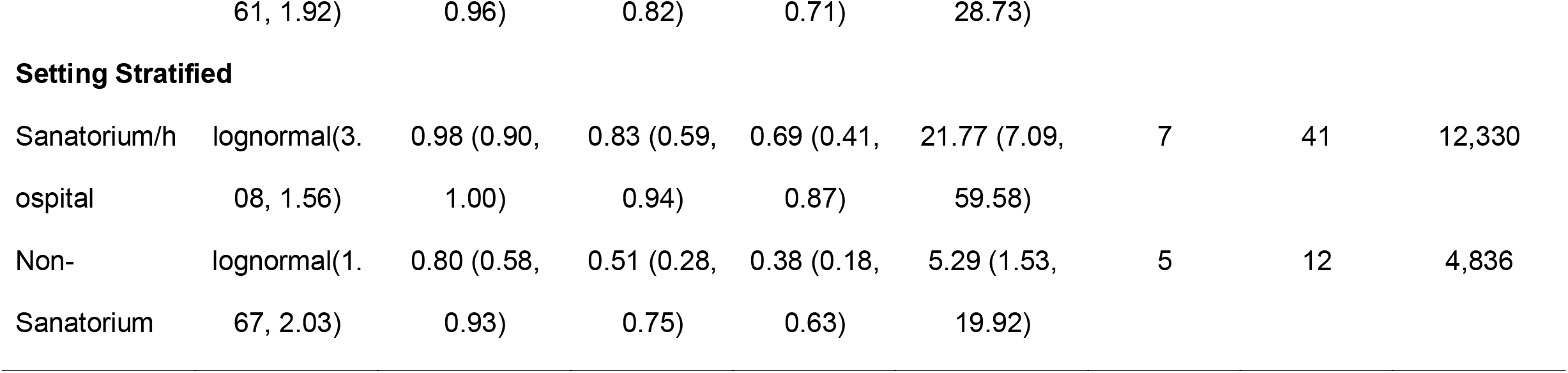
Survival analysis results TB-specific mortality with the combined and stratified models.

### Sanitoria results

Survival was higher in the sanatorium studies than in the non-sanatorium studies (Table 2, Figure 2, Figure S4). Only 38% (95% CI: 18%-63%) of non-sanitorium individuals survived to ten years compared to 69% (95% CI: 41%-87%) of sanitoria/hospitalized patients. There were more individuals with far advanced disease at baseline among those treated outside of sanitoria (43.0% versus 22.0%) and overall, there was a significant difference in disease severity between those treated in sanitoria compared to those treated elsewhere (p<0.001) (Table S7).

**Figure 2.**
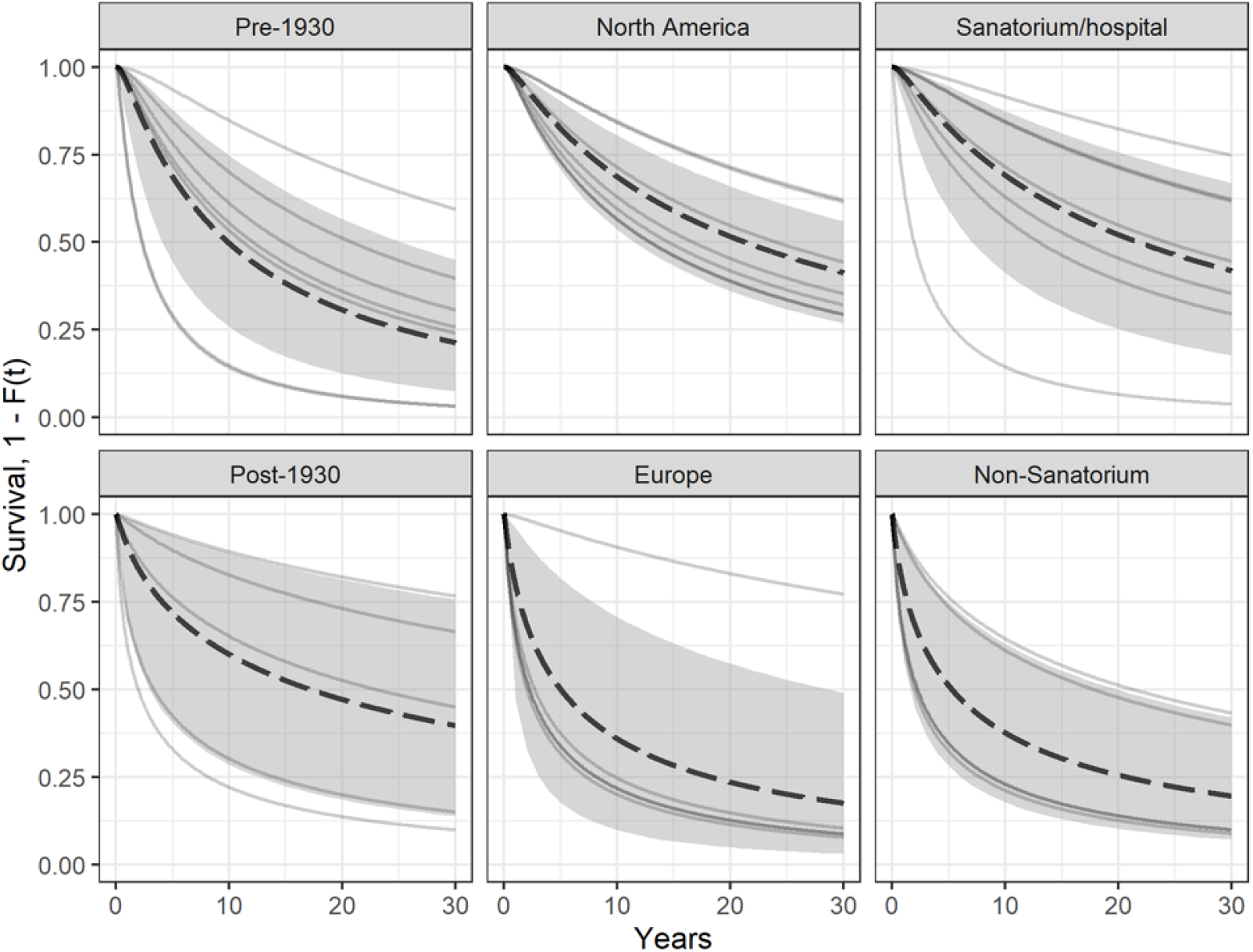
Survival curves for TB-specific mortality for the time period, study location, and treatment location stratified analyses. The thin, grey lines represent the specific survival curves for each study. The black dotted line represents the overall survival curve (the mean of the frailty distribution). The grey shaded area is the 95% credible interval for the overall survival curve.

### Mortality by time

In studies primarily enrolling individuals prior to 1930, ten-year survival was 50% (95% CI: 26%-75%), which was similar to survival estimated in studies primarily enrolling after 1930 [60% (95% CI: 29%-89%)]. Median TB survival time was nearly twice as long for studies conducted after 1930 (17.23 years, 95% CI: 2.98-129.05 versus 9.90 years, 95% CI: 4.10-25.24) (Table 2, Figures 2 and 3, Figure S5).

**Figure 3.**
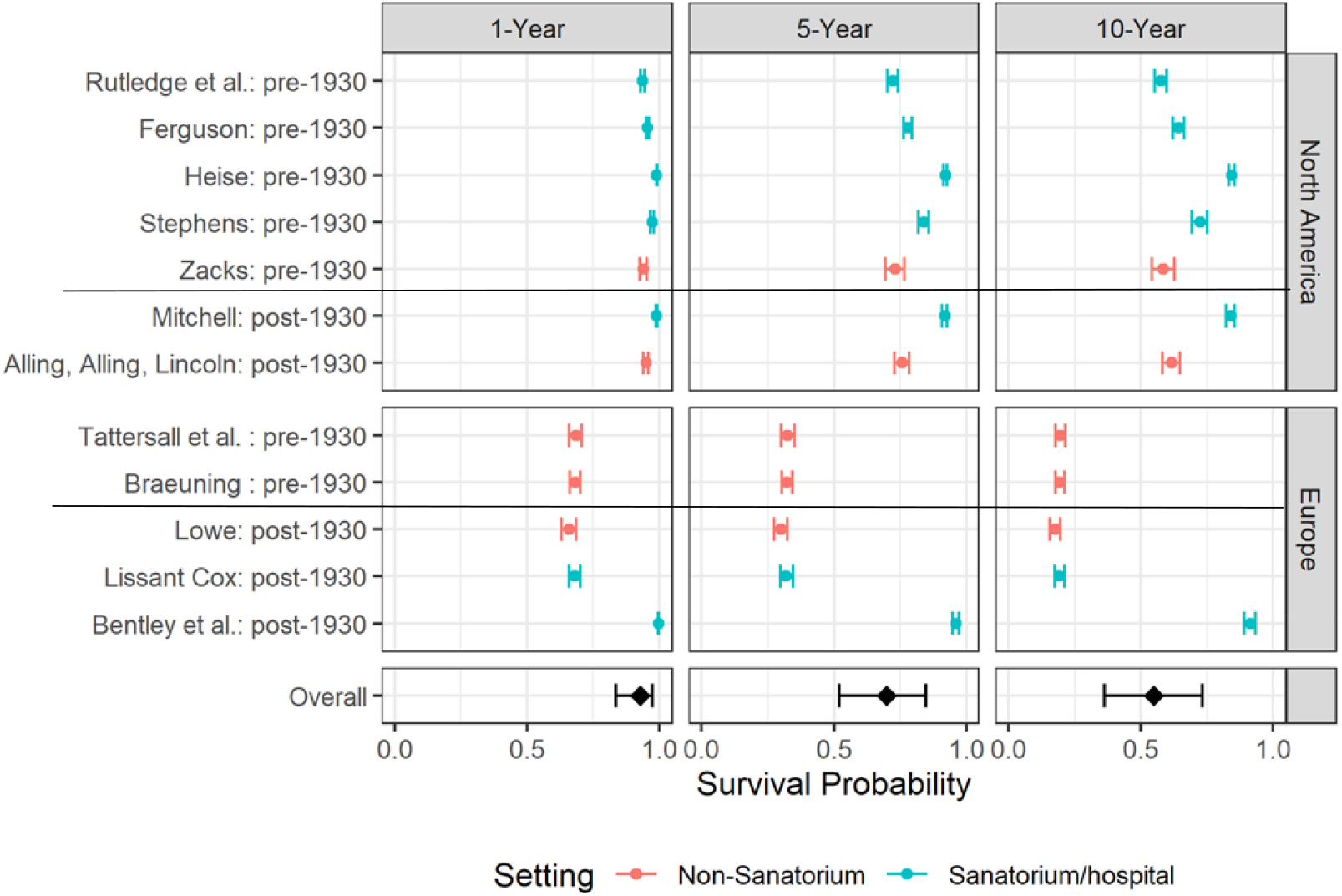
Forest plot of one, five, and ten-year survival probabilities for TB-specific mortality for the combined model. The studies are sorted by first year of enrollment (earliest to latest) with a horizontal line splitting the pre-1930 and post-1930 studies. The two panels for stratify the studies by geography and the dots and bars are colored by setting. The rows are the survival probabilities with 95% credible intervals for the different studies. The overall estimate of the survival probabilities is located at the bottom of the plot and represented with a diamond

### Mortality by geography

Overall mortality outcomes in North American studies compared to European studies were much better. Ten-year survival was estimated to be 69% in North America (95 CI: 54%-81%) compared to 36% in European studies (95% CI: 10%-71%). Median survival times were four-fold higher in North American studies: 21.33 years (95% CI: 11.54, 37.95) versus 5.02 (95% CI: 0.86, 28.73) (Table 2, Figure 2, Figure S6).

### Natural recovery data

The raw natural recovery data are summarized in the Supplement (Figures S6-S9). Consistent with findings from mortality data, overall trends in reported recovery rates were higher among individuals treated in sanitoria, post-1930 and from North America.

## DISCUSSION

Data from TB patients during the pre-chemotherapy era are a rich resource that can inform current TB management. However, our study illustrates that researchers should be mindful of the context when using these data. There are substantial differences in outcomes based on time, geography, and whether or not patients were in a sanatorium. Inferences from these data should take account of these potential biases and not view pre-chemotherapy era data as homogeneous.

Geographically, people in North American studies experienced better outcomes than those in Europe. This may be because North America was more shielded from the worst effects of the two world wars in the early 20^th^ century than Europe. Also, an increased proportion of the North American studies used data from sanatoria (6/8 studies versus 2/10 studies). Our results indicate that outcomes from sanatoria were better than in studies using community data. However, for studies that reported data on severity at presentation, we found that patients with less severe disease were more likely to spend time in a sanatorium, consistent with the hypothesis that sanatoria were less likely to admit moribund patients as they could not help them. Thus, providing an additional reason why data from North American studies could be biased towards improved outcomes, and illustrating a bias intrinsic to sanatoria data. Stadler et al. ^9^ also suggested that sanatoria preferentially treated people of higher socioeconomic status; even if their stay was paid, some people declined a sanatorium stay because they had to work to support family; and some people were excluded as “unsuited” for sanatoria. All of these factors are still seen today regarding the inequities of who does and does not receive TB care. We note that we did not observe significant differences in survival among studies enrolling individuals pre-1930 compared to those enrolling after 1930.

It is well documented that people with fewer resources, poor access to care, or live in political or social turmoil, such as war-torn areas, have poorer TB outcomes ^10–18^. Our results are consistent with this in that people without access to sanatoria, or living through wartime experienced poorer TB outcomes. Aggregating the pre-chemotherapy era data obscures this important heterogeneity. When making inferences from these data for modern day TB management, it might be necessary to identify specific relevant studies for analysis, depending on the research question.

A study strength is our focus on TB-specific mortality. Previous studies have looked at all-cause mortality, which would not only over-estimate TB-specific mortality, but also might suffer from different types of heterogeneities, thus obscuring differences in TB-specific mortality between groups ^2,3^. Also, we have used parametric survival modeling to estimate parameters that can be used to generate survival probabilities and parameterize modeling studies.

Our study has some limitations. The sanatorium discharge start time was used for the sanatoria studies with the average stay added (two of seven sanitoria studies) to this whereas it was notification/diagnosis for the non-sanatoria studies. Therefore, only individuals who survived their sanatorium stay were included. This may have contributed to improved survival in sanatorium studies. Asymptomatic patients were not included in our cohorts since they would not have presented to medical services.

Nonetheless, our analyses will have included nearly all symptomatic smear-positive patients. “A few” patients were reported to receive anti-TB drugs and therefore our estimated survivals might be overestimates. However, the availability of anti-TB drugs and types of regimens provided during our study period was limited and inferior to those used nowadays, so it is unlikely that this affected our results substantially. Lastly, our data do not account for the impact of HIV co-infection on TB survival and recovery, since they are from the pre-HIV era.

Since around 40% of people estimated to develop TB disease in 2021 were not diagnosed ^1^, results from the pre-chemotherapy era provide important insights that are relevant nowadays. These data are also critical for modeling studies. However, the context must be considered when using these data to ensure that we make suitable inferences for 21^st^ century TB management.

## Data Availability

All data and code are available on GitHub at https://github.com/sarahleavitt/TB_mortality.

## ACKNOWLEDGEMENTS

We acknowledge Margaret Zimmer for research assistance and Annelies Mesman for translation support. The results reported herein correspond to specific aims of grants R01GM122876 and R35GM141821 to investigator LFW, HEJ, and CRH, and R01GM122876-04S1 to CAR from the National Institute of General Medical Sciences of the National Institutes of Health. This work was also supported by grant K23 AI152930 from the National Institutes of Health to TCB and a Burroughs Wellcome Fund/American Society for Tropical Medicine and Hygiene Postdoctoral Fellowship in Tropical Infectious Diseases to TCB. The content is solely the responsibility of the authors and does not necessarily represent the official views of the National Institutes of Health.

## DATA AND CODING

We used R v4.0.2 with Bayesian models fit using JAGS with R2jags v0.6-1. All data and code are available on GitHub at https://github.com/sarahleavitt/TB_mortality.

