## Supplementary Material for "Survival of people with untreated tuberculosis: effects of time, geography and setting"

**Survival of people with untreated tuberculosis: effects of time, geography and context - Supplementary Material**

### **1. Outcomes of interest**

**Figure S1**

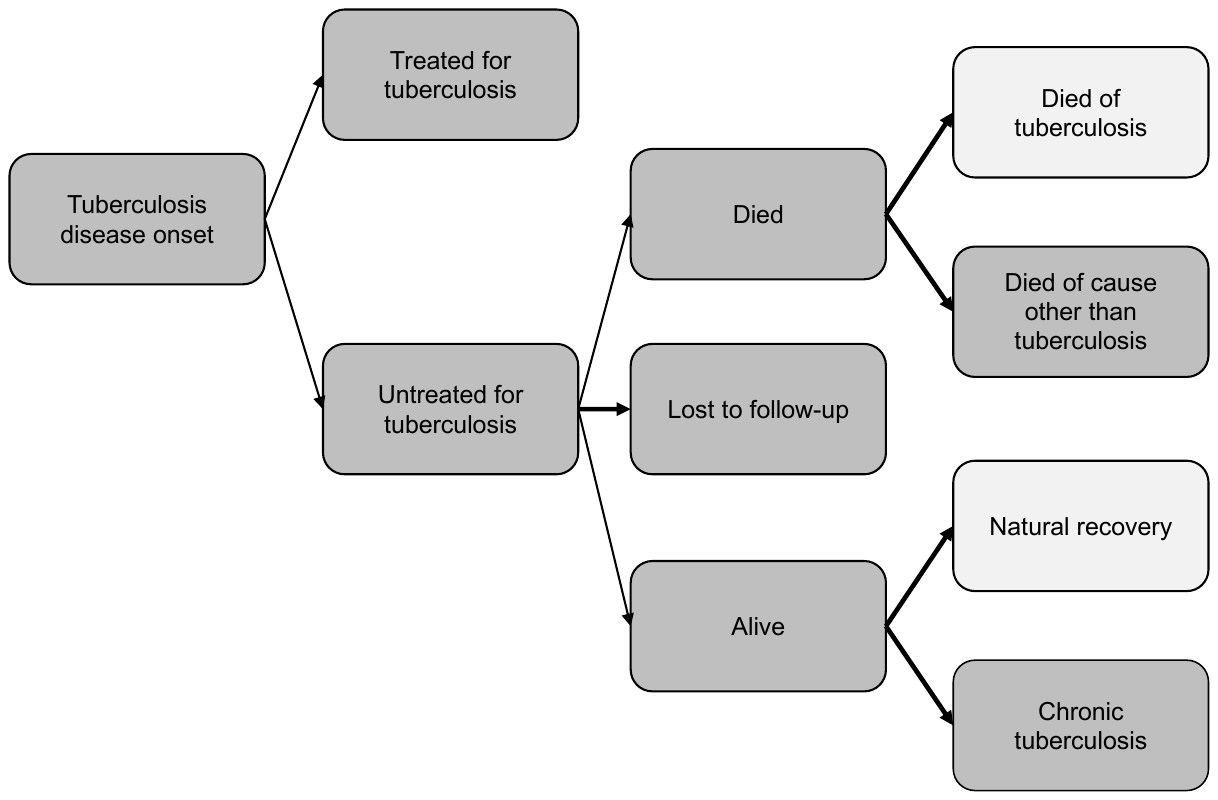

### **2. Review of papers included in Tiemersma et al.**

We reviewed all papers that had been included in Tiemersma et al.^1^, which used less strict inclusion criteria than we did in our study. Table S1 shows their included papers and whether or not we included the paper in our study, and reasons for exclusion where applicable.

**Table S1. Papers included in the study by Tiemersma et al.**^1^**, whether or not the study was included in our study, and reasons for exclusion, if applicable.** Note that citation numbers refer to the citation in Tiemersma et al. and include a link to that paper for ease of identifying the citation.

| Study | Include/Exclude |
| --- | --- |
| Hartley et al ^2^ | Could not locate full text. In fact. Tiemersma said: "We included the relevant studies that were not available to us in full text (Trail and Stockman (1931) [117], and Hartley, Wingfield and Burrows (1935) [118]), to the extent summarized by Berg [115]” |
| Sinding-Larsen ^3^ | Exclude, TB-specific mortality not presented (only all cause mortality presented) |
| Trail and Stockman | Could not locate full text. In fact. Tiemersma said: "We included the relevant studies that were not available to us in full text (Trail and Stockman (1931) [117], and Hartley, Wingfield and Burrows (1935) [118]), to the extent summarized by Berg [115]” |
| Backer ^4^ | Exclude, TB-specific mortality not presented (only all cause mortality presented) |
| Krebs ^5^ | Exclude, TB-specific mortality not presented (only all cause mortality presented) |
| Tattersall ^6^ | Included |
| Magnusson ^7^ | Exclude, TB-specific mortality not presented (only all cause mortality presented) |
| Rutledge and Crouch ^8^ | Included |
| Münchbach ^9^ | Included |
| Braeuning and Neisen ^10^ | Included |
| Braeuning and Neisen ^11^ | Exclude, overlapping patient sample with Braeuning and Neisen ^10^ |
| Griep ^12^ | Exclude, TB mortality presented but only present all cause mortality |
| Baart de la Faille ^13^ | Included |
| Buhl and Nyboe ^14^ | Exclude, TB-specific mortality not presented (only all cause mortality presented) |
| Lindhardt ^15^ | Exclude, TB-specific mortality not presented (only all cause mortality presented) |
| Berg ^16^ | Exclude, TB-specific mortality not presented (only all cause mortality presented) |
| Thompson ^17^ | Exclude, TB-specific mortality not presented (only all cause mortality presented) |
| National Tuberculosis Institute (NTI), Bangalore ^18^ | Exclude, TB treatment available for some individuals |
| Pamra et al ^19^ | Exclude, No TB mortality data |
| Drolet ^20^ | Exclude, population-level rates only |
| Braeuning ^21^ | Exclude, population-level rates only |
| Framingham Community Health & Tuberculosis Demonstration ^22–25^ | Exclude, TB-specific mortality not presented (only all cause mortality presented) |

### **3. Description of included studies**

**Table S2. Categorization of studies.**

| **Reference** | **Setting** | **Time** | **Geography** | **Analysis Type** |
| --- | --- | --- | --- | --- |
| Alling, 1954^26^, Alling, 1955^27^, Lincoln^28^ | Not sanatorium | Post-1930 | North America | Mortality, Natural recovery |
| Baart de la Faille^13^ | Sanatorium/hospital | Pre-1930 | Europe | Natural recovery |
| Bentley^29^ | Sanatorium/hospital | Post-1930 | Europe | Mortality |
| Braeuning^11^ | Not sanatorium | Pre-1930 | Europe | Mortality, Natural recovery |
| Ferguson^30^ | Sanatorium/hospital | Pre-1930 | North America | Mortality |
| Heise^31^ | Sanatorium/hospital | Pre-1930 | North America | Mortality |
| Holst^32^ | Not sanatorium | Pre-1930 | Europe | Natural recovery |
| Lissant Cox^33^ | Sanatorium/hospital | Post-1930 | Europe | Mortality |
| Lowe^34^ | Not sanatorium | Post-1930 | Europe | Mortality |
| Mitchell ^35^ | Sanatorium/hospital | Post-1930 | North America | Mortality |
| Munchbach^9^ | Sanatorium/hospital | Pre-1930 | Europe | Natural recovery |
| Rutledge^8^ | Sanatorium/hospital | Pre-1930 | North America | Mortality |
| Springett^36^ | Not sanatorium | Post-1930 | Europe | Natural recovery |
| Stadler^37^ | Not sanatorium | Pre-1930 | Europe | Natural recovery |
| Stephens^38^ | Sanatorium/hospital | Pre-1930 | North America | Mortality, Natural recovery |
| Tattersall ^6^ | Not sanatorium | Pre-1930 | Europe | Mortality |
| Wherrett[40] | Sanatorium/hospital | Pre-1930 | North America | Natural recovery |
| Zacks[41] | Not sanatorium | Pre-1930 | North America | Mortality |

### **4. Data extraction**

For Mitchell ^35^ we extracted the counts from a hand drawn figure resulting in errors in 5/40 of the time points. For each time point we expect the number of people in dead TB, dead other, natural recovery, chronic, and lost categories should add to the total sample size at that time point. In four time points the sum of the categories was greater than the total sample size. In those cases, we removed the excess people from the chronic category because in all four instances, the previous value of the chronic TB group was the same and the next value was a large decrease. The figure seemed to indicate a more steady and gradual decrease in this category suggesting that this was the most likely location of the extraction error. In ^35^ time point the sum of the categories was less than the total sample size. In that case, we added the missing people to the lost category because the previous value of the lost category was the same and the next value was a large increase. The figure seemed to indicate a more steady and gradual increase in this category suggesting that this was the most likely the location of the extraction error.

The Stephens paper ^38^ had three different tables; one for the three-year follow-up where the total sample size was the full cohort, one for five-year follow-up where the total sample size was those who could be followed for five years, and one for ten-year follow-up where the total sample size was those who could be followed for ten years. In each table, the total number of people who had died of any cause and of TB was recorded out of the total who could be followed to the time point of interest. This presented a challenge because the death counts were not strictly cumulative across the tables because the denominators changed. Instead, the death counts were overlapping to an unknown amount.

In order to estimate the number of people who died in each interval (0-3, 3-5, and 5-10 years) we first imputed the number of people who died at one time point who could not have been followed to the next time point (and therefore were excluded from that table). This was possible because the censoring in this study based strictly on hospital entry year and not patient loss to follow-up, there was no reason to suspect that those who could not be followed to a time point were any more or less likely to have died in that time point. This allowed us to treat the number of deaths as cumulative and then to find the number of people who died between each time point.

We estimated the number of people who died between each time point (Note: steps 3-5 are repeated for both deaths from TB and deaths from other causes):

1. We used the sample size at each time point in the original tables to determine how many people could not be followed to year five and to year 10.
2. We then found the proportion of people who could not be followed to years five and ten by dividing the number who could not be followed to that time point by the sample size at the previous time point.
3. We used that proportion to estimate the number of people who died in the previous time point that also could not be followed to the next time point (proportion * deaths in previous time point).
4. We then estimated the true total number of people who were dead at each time point as the number who are recorded as dead that time point (which included those who were dead at the previous time point and could be followed to that time point plus those who died between that time point and the one before) plus the estimate from step 3 of the number of people who were dead in the previous time point but could not be followed to this time point.
5. We estimated the number of people who died at each time point by subtracting the total people who were dead at the previous time point from those who were dead at that time point (step 4).
6. We estimated the number of people who were censored at each time point as the number who could not be followed to that time point from the one before (step 1) minus the estimate of those who could not be followed to that time point and died in the previous time point (step 3).
7. Finally, we calculated the new sample size each time point (includes those who were censored from the previous time point) as the sum of those who died of TB, died of other causes, experienced natural recovery, had chronic TB, or were censored.

**Table S3. Determination of TB-specific** **death and definitions of natural recovery from pre-treatment era cohorts (N=20** **publications, 18 studies)**

| **Reference** | **Determination of death** | **Natural recovery definition** |
| --- | --- | --- |
| Alling, 1954^26^, Alling, 1955 ^27^, Lincoln^28^ | NR | “Arrested: For ≥6 months prior to the [specified interval], the patient's sputum has been free of AFB and his serial chest roentgenograms have been compatible with stable disease. When more than a year has elapsed between clinic examinations and the chest roentgenograms show a stable lesion, the patient is continued in the arrested category throughout the interim.  Presumably arrested: The patient is known to be well and working full time but has not had a recent chest roentgenogram. The latest available roentgenograms are compatible with stable lesion. In the event that a lapse of more than a year without examination precedes evidence of reactivation, the patient is assigned to the presumably arrested category during the interim.” |
| Baart de la Faille ^13^ | N/A | Answering “good” in response to a questionnaire about health and ability to work. |
| Bentley^29^ | NR | “Remained well” |
| Braeuning ^11^ | Clinician determined cause of death | “Pulmonary lesions closed” |
| Ferguson[43] | NR | “The group well and working, as estimated by the patients themselves, corresponds more or less with the combined groups apparently cured, arrested, and apparently arrested, classified by physicians.” |
| Heise^31^ | NR | N/A |
| Holst^32^ | N/A | “Able to work” |
| Lissant Cox^33^ | NR | “Improved”, “not materially improved” |
| Lowe^34^ | NR | N/A |
| Mitchell ^35^ | Clinician-determined cause of death (no autopsy specified) | “…'well' represents the proportion of patients well and able to work in each year of observation” |
| Munchbach^9^ | NR | “Able to work” |
| Rutledge^8^ | Death certificate, annual questionnaire sent to household, clinician or county clerk reports | N/A |
| Springett^36^ | NR | NR |
| Stadler^37^ | NR | “Able to work” |
| Stephens^38^ | Questionnaire sent to household | “Able to work” |
| Tattersall^6^ | NR | N/A |
| Wherrett[40] | Reported by “'friends, family physicians, health officers, and nurses” […] “sanatorium review exams, Dept of Pensions and National Health, and the Travelling Consultants of the League” | “Apparently cured”, “arrested”, “apparently arrested”§ |
| Zacks[41] | NR | N/A |

**Abbreviations:** acid fast bacilli **(AFB);** applicable but not reported (NR); not applicable (N/A), (i.e. outcome not included in study)

§ Apparently cured, arrested, and apparently arrested are defined in the 1940 *Diagnostic Standards* as patients who have no constitutional symptoms; sputum negative for tubercle bacilli; lesions stationary, apparently healed according to X-ray without evidence of pulmonary cavity. These conditions must be fulfilled for two years under ordinary conditions of life (apparently cured), six months with the ability to have one hour’s walking exercise twice daily for the previous two months (arrested), and three months with the ability to have one hour’s walking exercise daily for the previous two months (apparently arrested).

**Table S4. Definitions of disease severity levels from studies which stratified by disease severity (N=11 publications, 9 studies)**

| **Reference** | **Minimal** | **Moderately advanced** | **Far advanced** |
| --- | --- | --- | --- |
| Alling, 1954^26^, Alling, 1955 ^27^, Lincoln^28^ | Stage of disease estimated from earliest chest roentgenogram by measuring the maximum diameter of all visible cavities, in accordance with the United States National Tuberculosis Association’s *Diagnostic Standard,* 1940 edition ^42^. Details from the *Diagnostic Standard* are quoted below: | | |
|  | Slight lesions without demonstrable excavation confined to a small part of one or both lungs. The total extent of the lesions, regardless of distribution, shall not exceed the equivalent of the volume of lung tissue which lies above the second chondrosternal junction and the spine of the fourth or body of the fifth thoracic vertebra on one side.  […] In pulmonary tuberculosis, the sputum contains tubercle bacilli frequently in early or minimal lesions if they are active, even when demonstrable evidence of cavity formation is absent. | One or both lungs may be involved. The total extent of the lesions may not exceed:   - Slight disseminated lesions which may extend through not more than the volume of 1 lung, or the equivalent of this in both lungs. - Dense and confluent lesions which may extend through not more than the equivalent of 1/3 the volume of one lung. - Any gradation within the above limits.   Total diameter of cavities, if present, estimated not to exceed 4 cm. | Lesions more extensive than Moderately advanced. |
| Braeuning^11^ | Extend of lung involvement was assess from chest x-rays as three levels defined as follows and used in the minimal, moderately advanced and far advanced categories below: (1) Tip or width of an anterior rib margin plus intercostal space, (2) Tip to the hilum or three intercostal spaces wide, (3) More than (2). | | |
|  | Unilateral or bilateral lung involvement level (1) without cavities | Unilateral lung involvement level (1) with cavities, unilateral lung involvement level (2) with or without cavities, bilateral lung involvement level (1) with cavities | Unilateral lung involvement level (3) with or without cavities, bilateral lung involvement level (2) with or without cavities, bilateral lung involvement with at least one lung at level (3), with or without cavities |
| Heise^31^ | Minimal TB, moderately advanced TB and far advanced TB as classified by “X-ray examination of the lungs and whose condition has been classified according to the method of the United States National Tuberculosis Association adopted in 1930 and published by that association in January 1931 in a pamphlet termed *Diagnostic Standards.*”  Original 1931 source document could not be located to extract definitions. | | |
| Lissant Cox^33^ | TB plus I (early):  Tubercle bacilli have been demonstrated in the sputum, pleural fluid, faeces, etc.  Cases with slight constitutional disturbance (if any); e.g., there should not be marked acceleration of pulse nor elevation of temperature except of very transient duration; gastrointestinal disturbance or emaciation, if present, should not be excessive. The obvious physical signs should be of very limited extent, as follows: either present in one lobe only, and in the case of an apical lesion of one upper lobe, not extending below the second rib in front or not exceeding an equivalent area in any one lobe; or where the physical signs are present in more than one lobe, they should be limited to the apices of the upper lobes, and should not extent below the clavicle and the spine of the scapula. No complication (tuberculous or other) of prognostic gravity should be present. A small area of dry pleurisy should not exclude a case from this group. | TB plus II (intermediate):  Tubercle bacilli have been demonstrated in the sputum, pleural fluid, faeces, etc.  All cases which cannot be places in TB plus I and TB plus III. | TB plus III (advanced):  Tubercle bacilli have been demonstrated in the sputum, pleural fluid, faeces, etc.  Cases with profound systemic disturbance or constitutional deterioration, with marked impairment of function, either local or general, and with little or no prospect of recovering. All cases with grave complications (e.g., diabetes, tuberculosis of intestine, etc.), whether those complications are tuberculous or not, should be classified in this group. |
| Mitchell^35^ | N/A | No definitions provided  Treatment period would fall into the United States National Tuberculosis Association’s *Diagnostic Standard,* 1931, 1935, and 1938 editions.  Original source documents could not be located to extract definitions. | |
| Rutledge^8^ | Classification during the study period would have been dictated by ^43^:   - Report of Committee on Clinical Nomenclature, Transactions of National Association for the Study and Prevention of Tuberculosis, 1905, 1906, 1907, and 1908 - Report of Committee on Nomenclature of the American Sanatorium Association, Transactions of National Association for the Study and Prevention of Tuberculosis, 1912 - Report of Committee of Seven, Transactions of National Association for the Study and Prevention of Tuberculosis, 1913   Original source documents could not be located to extract definitions. Details from the publication are quoted below: | | |
|  | Incipient TB: “cases where a diagnosis of tuberculosis is practically certain. In these cases the diagnosis is made from history […] the sputum was free from bacilli, but where a history of hemorrhage, pleurisy with effusion, rectal fistula, or the history of tuberculosis in some other part of the body practically clinches the diagnosis.” | Moderately advanced TB: “diagnosed from the lung findings, regardless of sputum.” | Far advanced TB: “diagnosed from the lung findings, regardless of sputum.” |
| Stephens^38^ | No definitions provided  Classification during the study period would have been dictated by the United States National Tuberculosis Association’s *Diagnostic Standard,* 1920, 1922, 1926, 1928, 1931, and 1938 editions.  Original source documents for 1920 and 1922 were located ^44,45^, as quoted below. Source documents for other years could not be located to extract definitions. | | |
|  | 1920: *“Incipient TB”* Slight infiltration limited to the apex of one or both lungs or a small part of one lobe. No tuberculous complications. Constitutional symptoms (including gastric or intestinal disturbance or rapid loss of weight) may be slight or absent. There may be slight or no elevation of temperature or acceleration of pulse at any time during the 24 hours. The expectoration is usually small in amount of absent. Tubercle bacilli may be present or absent.  1922: *Same as 1920* | 1920: Marked infiltration more extensive than under incipient, with little or no evidence of cavity formation. No serious tuberculous complications and no marked impairment of function, either local or constitutional.  1922: *Same as 1920* | 1920: Extensive localized infiltration or consolidation in one or more lobes or disseminated areas of cavity formation. Serious tuberculous complications, with marked impairment of function, local, or constitutional .  1922: *Same as 1920* |
| Tattersall^6^ | Stage I, Stage II, Stage III. The text details “…the definition of the stages of the disease cannot be precisely stated, and must be left in relative terms of ‘early,’ ‘moderate,’ and advanced.’ The reason for this is that the cases in the present series are classified in the dispensary index as Stage I, Stage II, Stage III. Since the introduction of the Ministry of Health classification this scheme appears to have been used, but prior to this, however, no definition of the three categories can be found, though it is evident from the available records that the division is very similar to the Ministry of Health definition.” | | |

**Abbreviations:** not applicable (N/A), (i.e. severity level not included in study)

### **5. Statistical methods**

We provide more detail on the statistical methods used in the analyses. In brief, we used a parametric survival model to estimate the time to death from TB

For each individual ($i=1,\ldots, n$), we observe the following information: $\left( L_{i}, R_{i}, \delta_{i} \right)$ where $L_{i}$ is the left time point of the interval in which the individual died, or the time of censoring; $R_{i}$ is the right time point of the interval; and $\delta_{i}$ is equal to 1 if the individual died, and 0 if the individual was censored. We then use these data to estimate the probability density function (pdf) of the survival time, $f(t)$

$$L=\prod_{i=1}^{n} \left[ S\left( L_{i} \right)-S(R_{i})] \right]^{\delta_{i}}\left[ S(L_{i}) \right]^{1-\delta_{i}}$$

where $S\left( t \right)=P(T>t)$ is the survival function. For all analyses, we assumed that $f(t)$ followed a lognormal distribution with parameters meanlog ($\mu$) and sdlog ($\sigma$). Initially, we explored Weibull, exponential, and log-logistic distributions. All resulted in similar outcomes but the lognormal was chosen because it had the lowest AIC in numerous individual study analyses.

We used a Bayesian framework to obtain posterior estimates of the parameters. We incorporated a frailty term allowing each study to have its own meanlog parameter $u_{j},$ which were all assumed to come from a common distribution $N(u_{all}, \theta)$ with $\theta$ representing the frailty variance. Here, the overall estimate of the meanlog parameter was $u_{all}$ and $\sigma$ was assumed to be constant across all studies levels.

Non-informative priors were used for all initial parameters ($\theta$, $\sigma$, $u_{all}$The model structures is in Supplementary Figure S2.

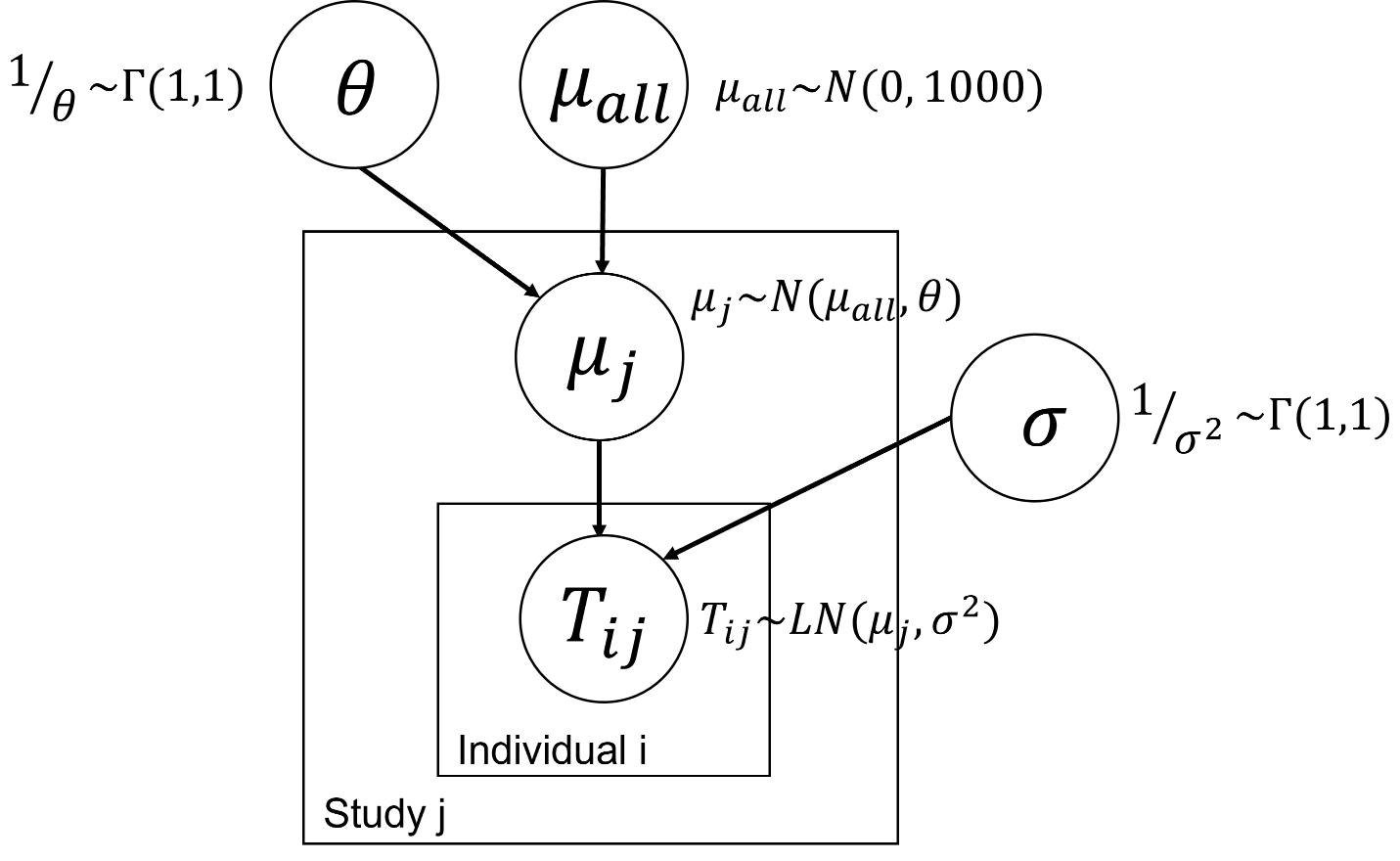

**Figure S2. Model diagram.** Here $T_{ij}$ is the survival time for individual $i$ (inner box) in study $j$ (outer box) which is assumed to follow a lognormal distribution with meanlog $\mu_{j}$ and sdlog $\sigma$. The meanlog parameters are unique to each study but are assumed to come from a normal distribution with mean $\mu_{all}$ and variance $\theta$ (the frailty variance). The parameters $\mu\_all$ and $\sigma$ can be used to find the overall lognormal survival distribution. All initial parameters: $\sigma$, $\mu_{all}$, and $\theta$ are initialized with non-informative priors as specified outside of the boxes.

**6. Additional results**

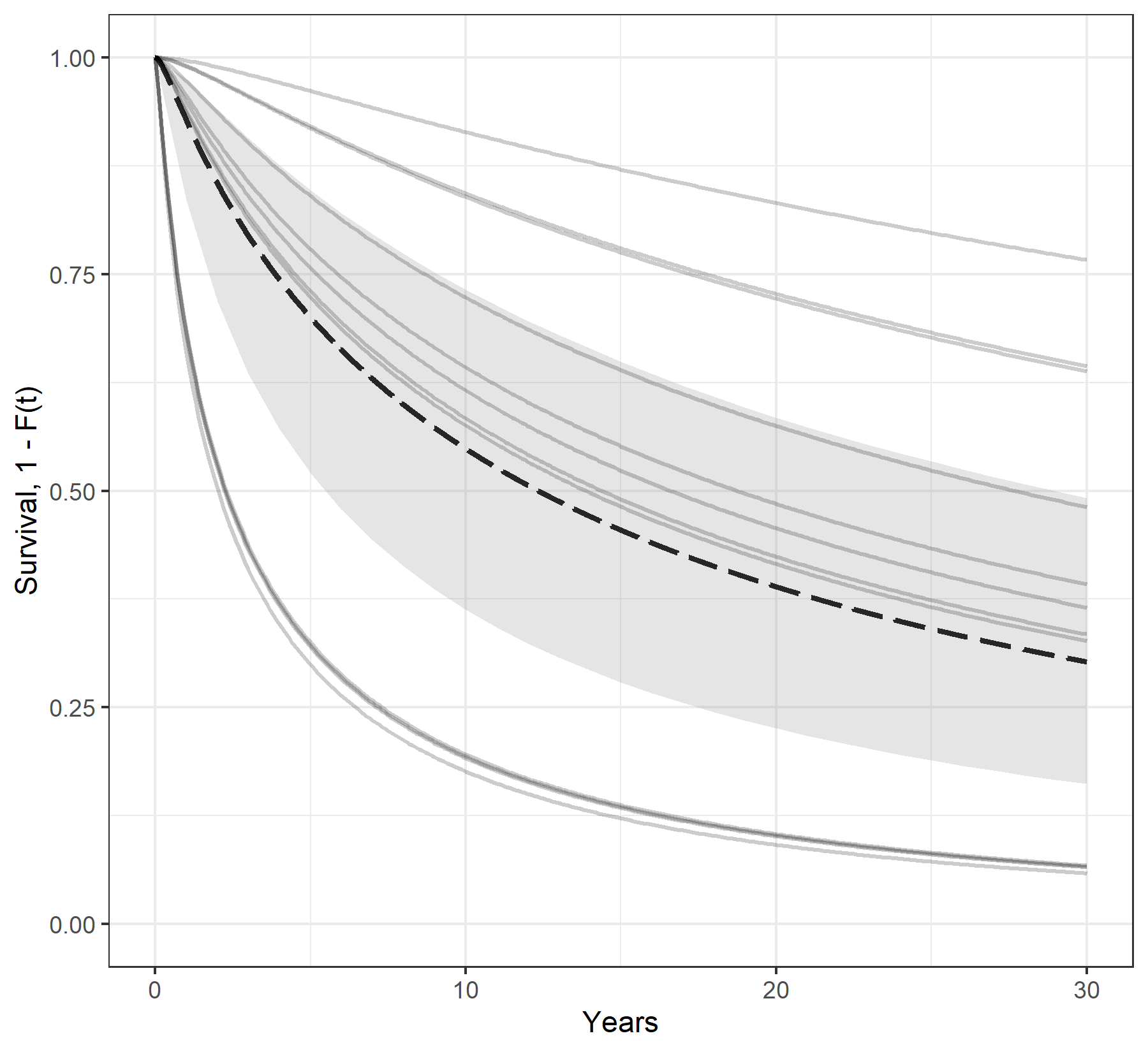

**Figure S3. Survival curves for TB-specific mortality for the combined analysis.** The thin, grey lines represent the specific survival curves for each study. The black dotted line represents the overall survival curve (the mean of the frailty distribution). The grey shaded area is the 95% credible interval for the overall survival curve.

**Table S5. Frailty variance for the TB survival analysis**

| **Model** | **Frailty Variance** |
| --- | --- |
| Combined | 1.92 |
| Pre-1930 | 1.27 |
| Post-1930 | 3.13 |
| North America | 0.57 |
| Europe | 2.78 |
| Sanatorium/hospital | 1.57 |
| Non-sanatorium | 1.53 |

**Table S6. Disease severity distribution for studies which stratify by severity**

| **Reference** | **Setting** | **Minimal** | **Moderately Advanced** | **Far Advanced** |
| --- | --- | --- | --- | --- |
| Alling, Alling, Lincoln | Non-sanatorium | 448 (50.7%) | 211 (23.9%) | 224 (25.4%) |
| Tattersall et al. | Non-sanatorium | 126 (12%) | 580 (55.4%) | 340 (32.5%) |
| Braeuning | Non-sanatorium | 4 (0.5%) | 155 (20.8%) | 587 (78.7%) |
| Mitchell | Sanatorium/hospital | 0 (0%) | 1206 (80.2%) | 298 (19.8%) |
| Rutledge et al. | Sanatorium/hospital | 173 (12%) | 591 (41.1%) | 674 (46.9%) |
| Stephens | Sanatorium/hospital | 334 (33.9%) | 458 (46.5%) | 192 (19.5%) |
| Lissant Cox | Sanatorium/hospital | 103 (12.5%) | 573 (69.7%) | 146 (17.8%) |
| Heise | Sanatorium/hospital | 708 (21%) | 2192 (64.9%) | 478 (14.2%) |

**Table S7.** **Disease severity distribution for sanatorium/hospital studies verses non-sanatorium studies at baseline.** Using a chi-square test of association, p<0.001, indicating a strong association between disease severity and treatment location.

| **Treatment Location** | **Minimal** | **Moderately Advanced** | **Far Advanced** |
| --- | --- | --- | --- |
| Non-sanatorium^a^ | 578 (21.6%) | 946 (35.4%) | 1151 (43.0%) |
| Sanatorium/hospital^b^ | 1318 (16.2%) | 5020 (61.8%) | 1788 (22.0%) |

^a^ Includes 3 studies

^b^ Includes 5 studies

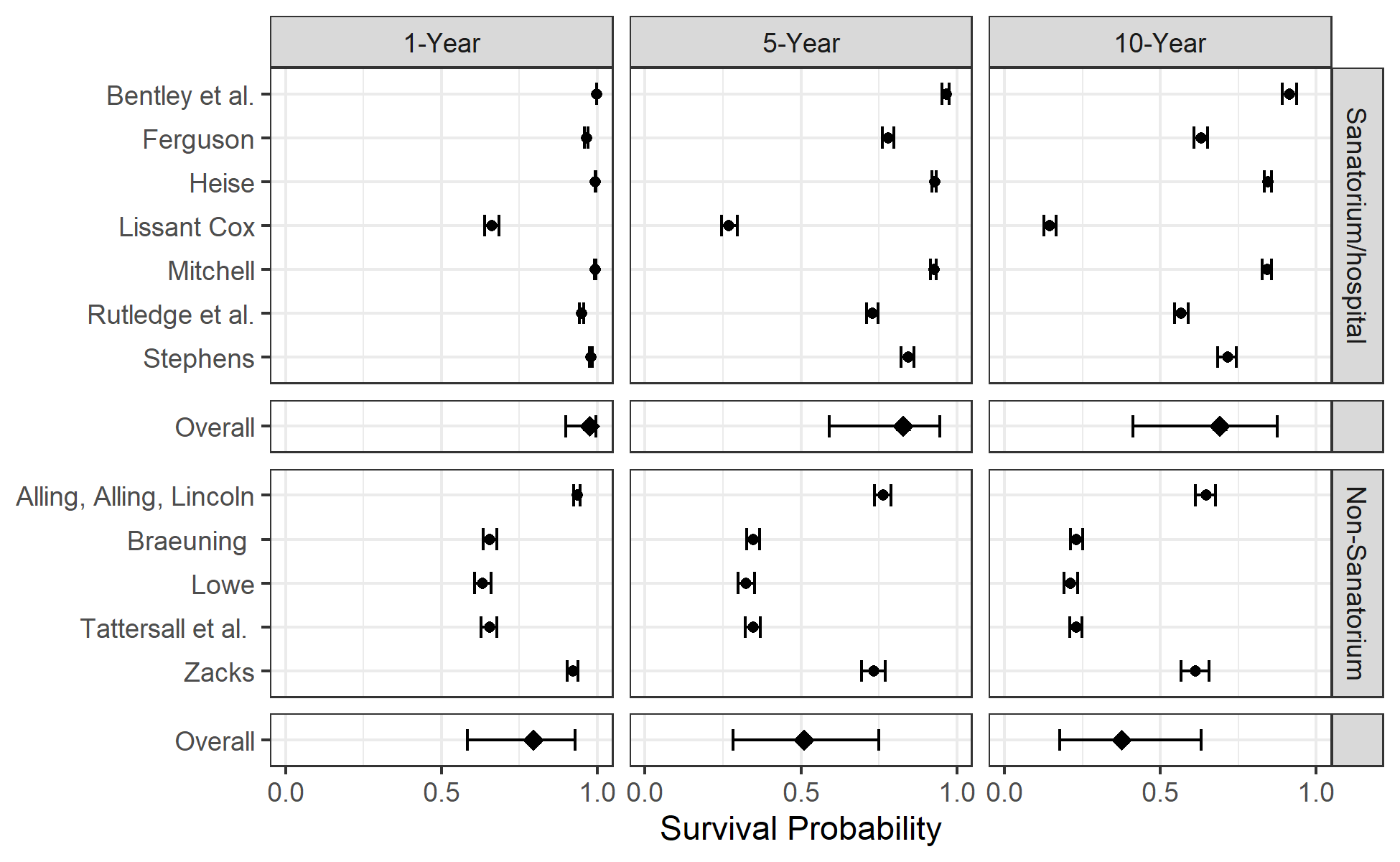

**Figure S4. Forest plot of one, five, and ten-year survival probabilities for TB-specific mortality for setting stratified model.** The rows are the survival probabilities with 95% credible intervals for the different studies. The overall estimates of the survival probabilities for each stratum are located at the bottom of the plot and represented with a diamond.

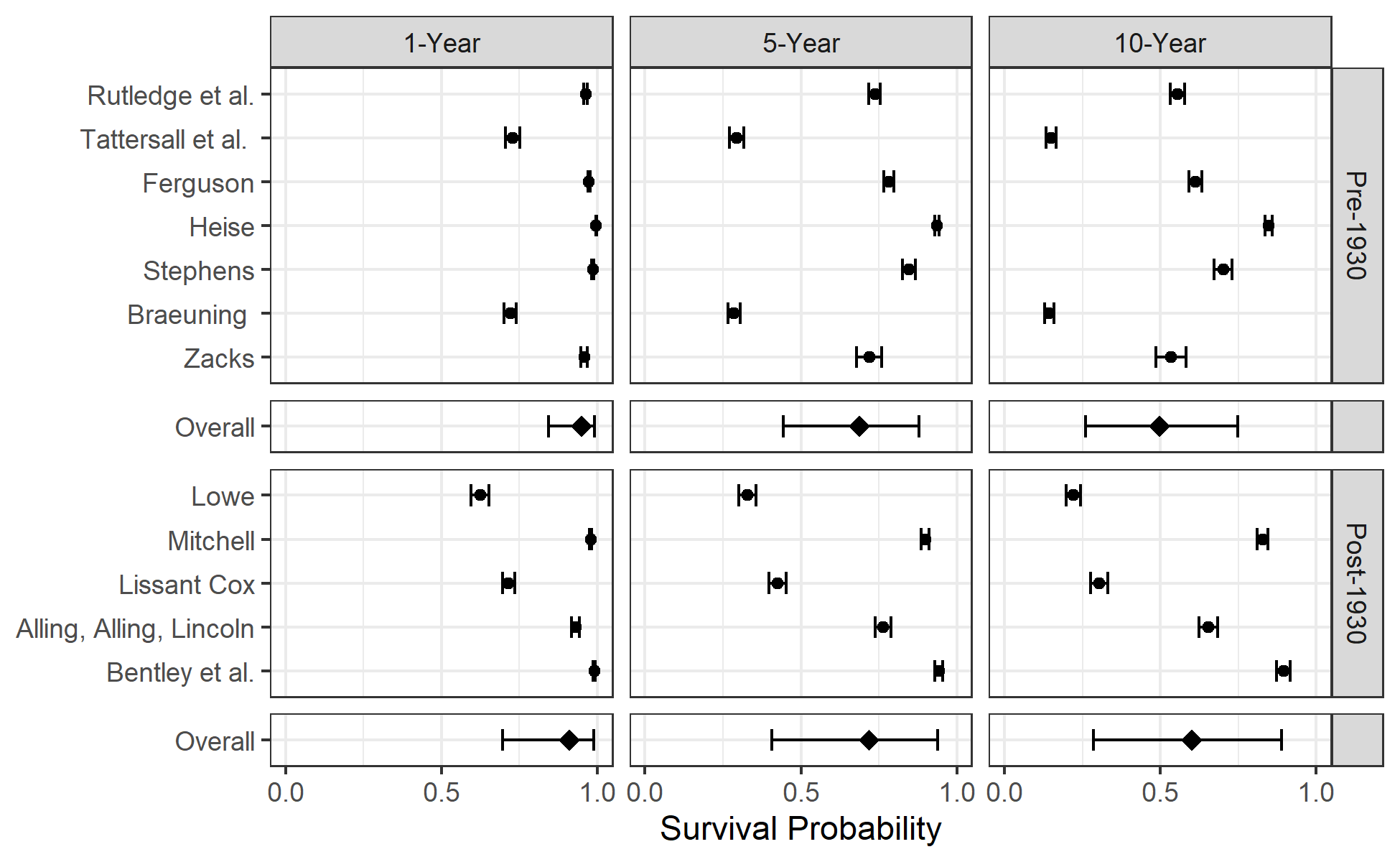

**Figure S5 Forest plot of one, five, and ten-year survival probabilities for TB-specific mortality for time period stratified model.** The rows are the survival probabilities with 95% credible intervals for the different studies and are ordered by first year of enrollment (earliest to latest). The overall estimates of the survival probabilities for each stratum are located at the bottom of the plot and represented with a diamond.

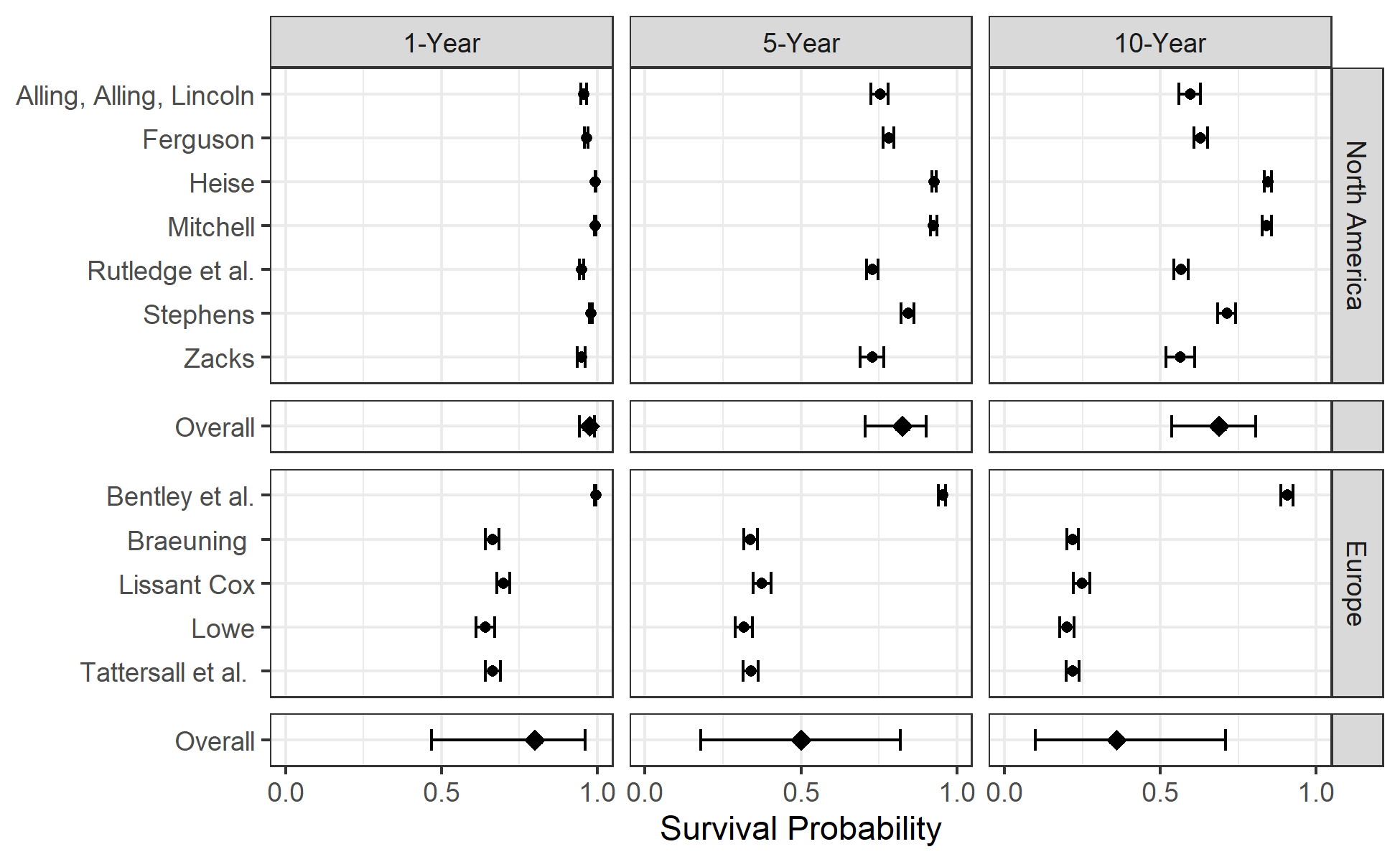

**Figure S6. Forest plot of one, five, and ten-year survival probabilities for TB-specific mortality for geography stratified model.** The rows are the survival probabilities with 95% credible intervals for the different studies. The overall estimates of the survival probabilities for each stratum are located at the bottom of the plot and represented with a diamond

**Natural Recovery Data**

We summarize the data from all papers with natural recovery information. In all 13 studies reported data on natural recovery sufficiently to summarize. When a cohort was followed longitudinally with loss to follow-up tracked, we connect the data with lines. Otherwise, the data are shown as individual points.

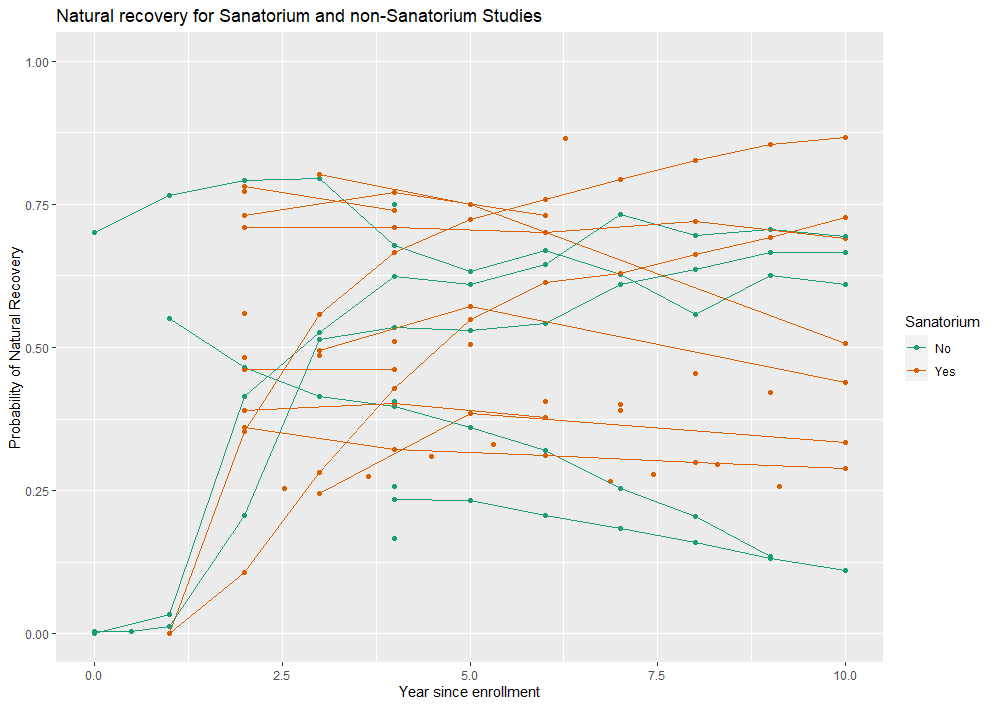

**Figure S7. Natural recovery data by setting of study (sanatoria versus community cohort).** Points that are connected by lines represent cohorts with longitudinal follow-up and lost to follow-up tracked.

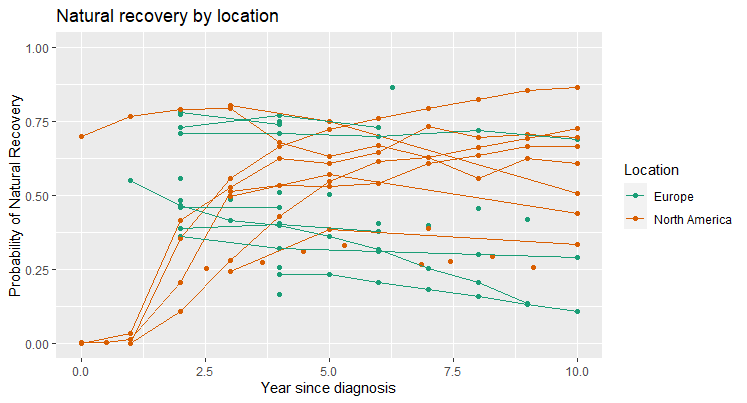

**Figure S8. Natural recovery data by location of study (Europe versus North America).** Points that are connected by lines represent cohorts with longitudinal follow-up and lost to follow-up tracked.

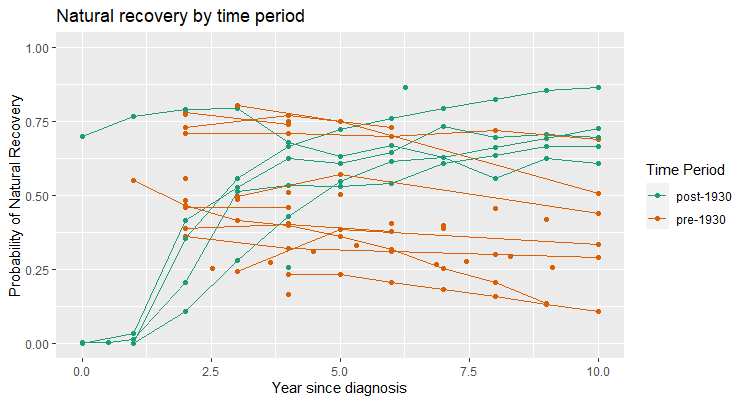

**Figure S9. Natural recovery data by time period of study (pre- versus post-1930).**

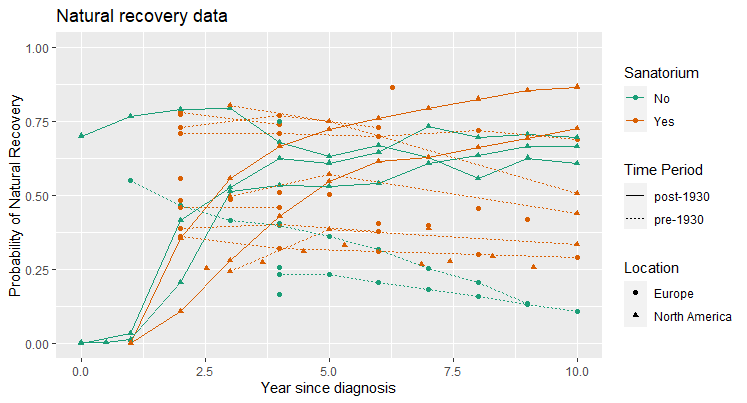

**Figure S10. Natural recovery data by time period, location and setting of study.**

**Model Diagnostics**

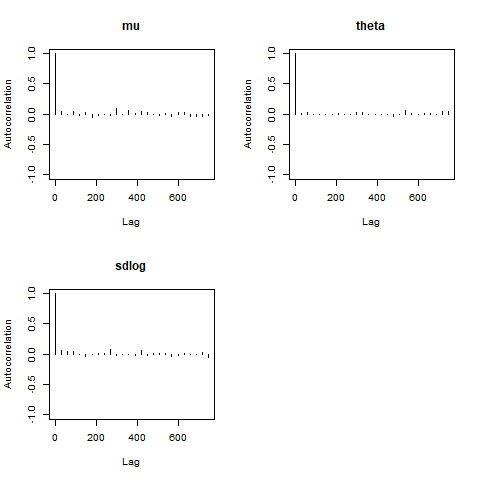

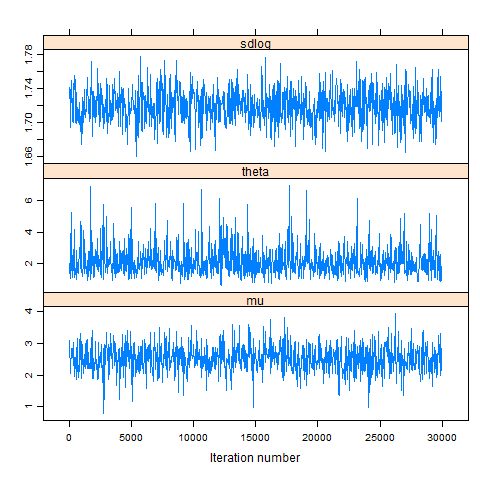

**Figure S11. Model diagnostics for combined model of TB mortality**

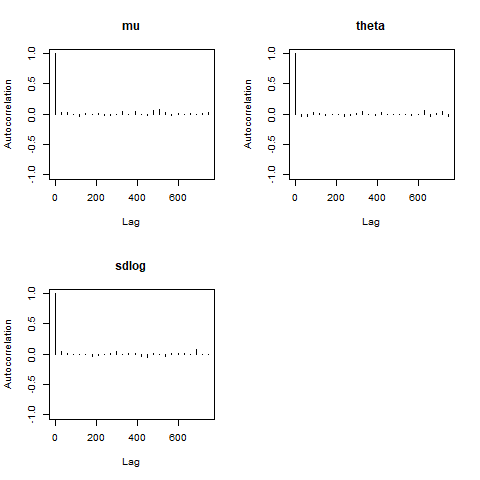

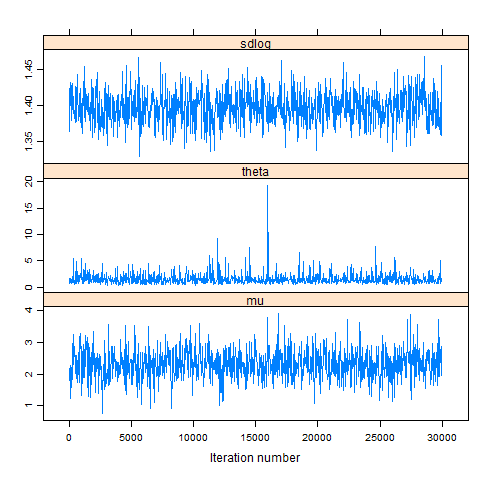

**Figure S12. Model diagnostics for TB mortality of pre-1930 studies**

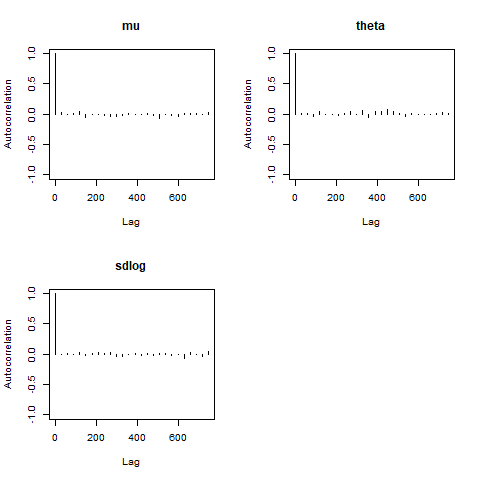

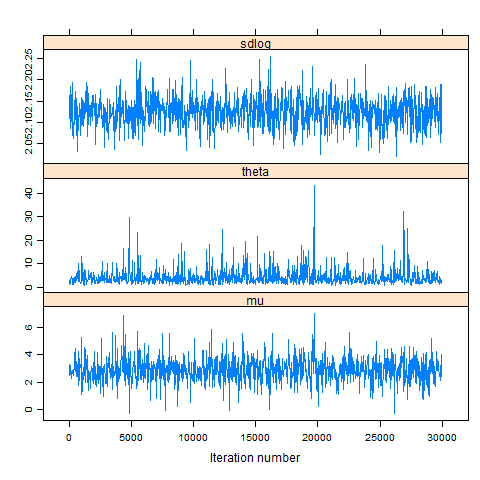

**Figure S13. Model diagnostics for TB mortality of post-1930 studies**

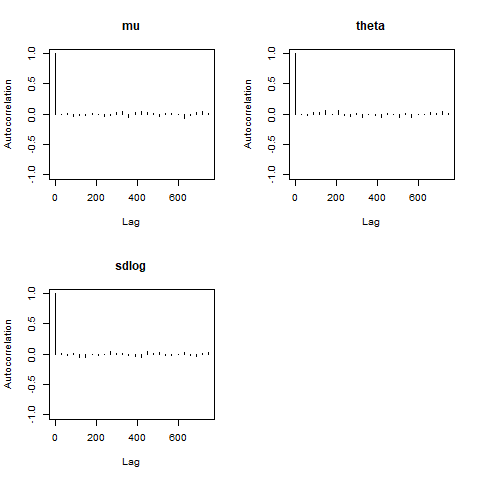

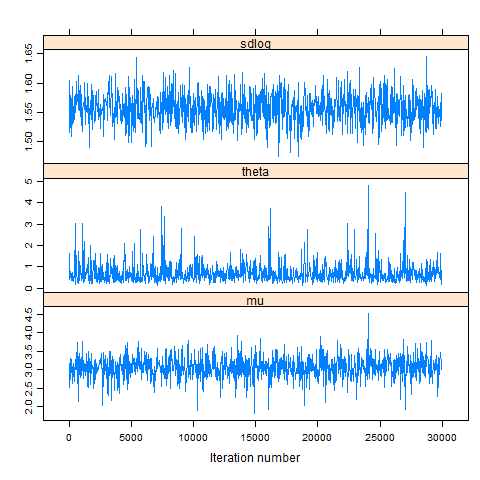

**Figure S14. Model diagnostics for TB mortality of North American studies**

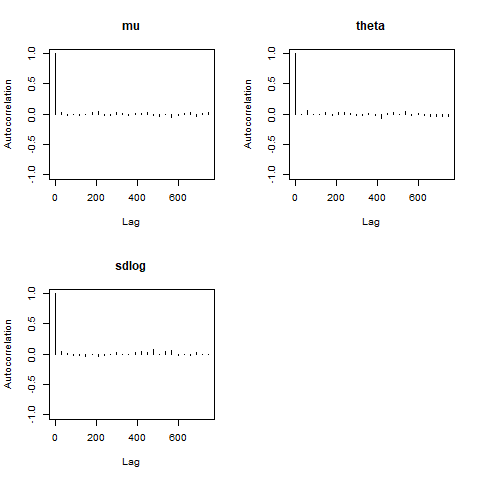

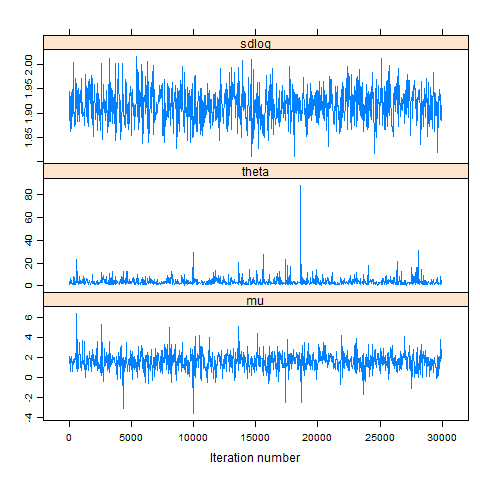

**Figure S15. Model diagnostics for TB mortality of European studies**

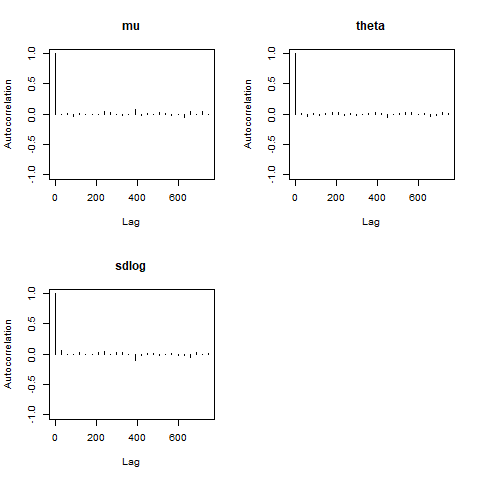

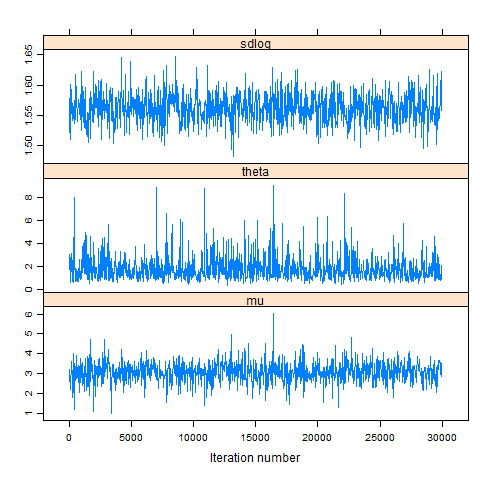

**Figure S16. Model diagnostics for TB mortality of Sanatorium/hospital studies**

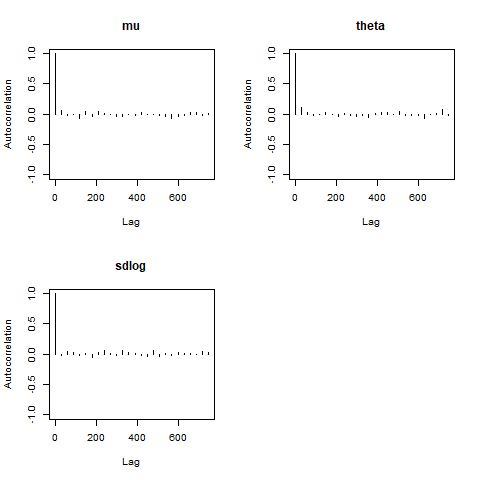

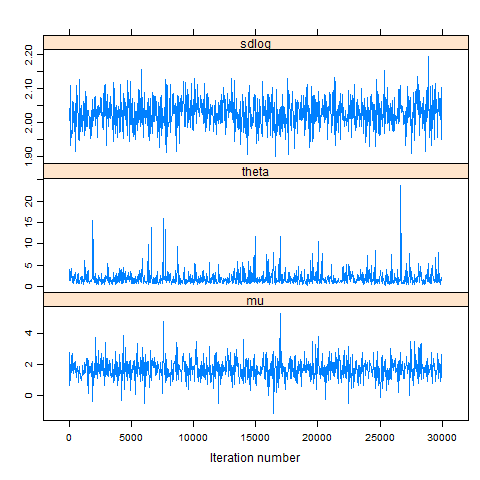

**Figure S17. Model diagnostics for TB mortality of Non-sanatorium studies**

43.	History of Diagnostic Standards and Classification of Tuberculosis of the National Tuberculosis Association. *Am Rev Tuberc*. 65(4):494-503.

44.	*Diagnostic Standards and Classification of Tuberculosis*.; 1920. https://catalog.hathitrust.org/Record/000053126

45.	Association NT. *Diagnostic Standards and Classification of Tuberculosis.*; 1922.
